# Vaccine nationalism and the dynamics and control of SARS-CoV-2

**DOI:** 10.1101/2021.06.02.21258229

**Authors:** Caroline E. Wagner, Chadi M. Saad-Roy, Sinead E. Morris, Rachel E. Baker, Michael J. Mina, Jeremy Farrar, Edward C. Holmes, Oliver G. Pybus, Andrea L. Graham, Ezekiel J. Emanuel, Simon A. Levin, C. Jessica E. Metcalf, Bryan T. Grenfell

**Affiliations:** Department of Bioengineering, McGill University, Montreal QC H3A 0C3, Canada; Lewis-Sigler Institute for Integrative Genomics, Princeton University, Princeton NJ 08540, USA; Department of Pathology and Cell Biology, Columbia University Medical Center, New York NY 10032, USA; Department of Ecology and Evolutionary Biology, Princeton University, Princeton NJ 08540, USA; Princeton High Meadows Environmental Institute, Princeton University, Princeton NJ 08540, USA; Departments of Epidemiology and Immunology and Infectious Diseases, Harvard T. H. Chan School of Public Health, Boston MA 02115, USA; The Wellcome Trust, London, UK; Marie Bashir Institute for Infectious Diseases and Biosecurity, School of Life and Environmental Sciences and School of Medical Sciences, The University of Sydney, Sydney, NSW, Australia; Department of Zoology, University of Oxford, Oxford, UK; Department of Medical Ethics and Health Policy, Perelman School of Medicine, University of Pennsylvania, Philadelphia, PA, USA; Princeton School of Public and International Affairs, Princeton University, Princeton NJ 08540, USA

## Abstract

Vaccines provide powerful tools to mitigate the enormous public health and economic costs that the ongoing SARS-CoV-2 pandemic continues to exert globally, yet vaccine distribution remains unequal between countries. To examine the potential epidemiological and evolutionary impacts of ‘vaccine nationalism’, we extend previous models to include simple scenarios of stockpiling. In general, we find that stockpiling vaccines by countries with high availability leads to large increases in infections in countries with low vaccine availability, the magnitude of which depends on the strength and duration of natural and vaccinal immunity. Additionally, a number of subtleties arise when the populations and transmission rates in each country differ depending on evolutionary assumptions and vaccine availability. Furthermore, the movement of infected individuals between countries combined with the possibility of increases in viral transmissibility may greatly magnify local and combined infection numbers, suggesting that countries with high vaccine availability must invest in surveillance strategies to prevent case importation. Dose-sharing is likely a high-return strategy because equitable allocation brings non-linear benefits and also alleviates costs of surveillance (e.g. border testing, genomic surveillance) in settings where doses are sufficient to maintain cases at low numbers. Across a range of immunological scenarios, we find that vaccine sharing is also a powerful tool to decrease the potential for antigenic evolution, especially if infections after the waning of natural immunity contribute most to evolutionary potential. Overall, our results stress the importance of equitable global vaccine distribution.

---

The SARS-CoV-2 pandemic has led to more than 100 million infections and nearly 3 million fatalities to date (*1*). Effective vaccines (e.g. (*2–4*)) have now been approved and are actively being deployed, but numerous important questions remain. Eventually, community immunity may be attained through the deployment of vaccines; however if and when this occurs will be contingent on the characteristics of natural and vaccinal immunity (*5–7*) in conjunction with SARS-CoV-2 evolutionary potential.

Due to strong public and political pressures and fear of waning immunity, some countries with high vaccine availability are currently resorting to ‘vaccine nationalism’: stockpiling vaccines to prioritize rapid access to their citizenry (*8*). Recently, the World Health Organization recognized that delayed access to vaccines in countries with low vaccine availability may lead to more evolutionary potential for immune escape (*9*). Indeed, at the time of writing, 86 and 91 doses per 100 individuals have been administered in the United States and United Kingdom, respectively, while an average of 14 and 2.1 doses per 100 individuals have been administered in India and across Africa, respectively (*10*). The emergence of future variants capable of evading natural or vaccinal immune responses could threaten containment efforts globally. These concepts underlie the development of a number of policy tools, including the existing COVAX initiative. Furthermore, to ensure that vaccine distribution is ethically-sound and equitable, the “Fair Priority Model” has been proposed (*11–13*) as a potential replacement to the currently planned proportional allocation (by population size) from COVAX.

Prior work exploring optimal prophylactic vaccine allocation for minimizing the final epidemic size of a fully immunizing infection (i.e. one that can be modeled using a susceptible-infected-recovered (SIR) framework) found that when interaction between communities (or countries) is considered, equal vaccine distribution is increasingly advantageous in terms of minimizing case numbers (*14*). Modeling studies have also shown that coordinated influenza vaccine sharing would reduce the financial and infection burden of influenza outbreaks globally (*15*). Similar problems related to optimizing vaccine allocation have also been explored in networks with community structure (*16*) as well as in the face of economic constraints (*17*) and coalition formation (*18*), and for SARS-CoV-2 with age- (and contact-) structure (*19, 20*). We have recently shown that the strength and duration of immunity elicited following infection or one or two doses of a vaccine will have a crucial impact on the medium-term epidemiological and potential evolutionary outcomes (*5, 6*). Here we extend these analyses to address potential epidemiological and evolutionary consequences of policies of vaccine nationalism or equitable access for a range of assumptions regarding the robustness of host immune responses. In reality, vaccine distribution is a public goods problem, and the optimal “global” allocation projected based on evolutionary and immunological uncertainties may differ from national optima due to the actual economic landscape of each country. In all cases, however, the reduction in the potential for novel strains to arise associated with minimizing the global infection burden is likely critical.

We consider a trans-national extension of our model comprised of two countries with possibly different population sizes and seasonal transmission patterns. One country, the high access region (HAR), chooses to allocate a fraction *f* of the total vaccine supply to the low access region (LAR). The underlying immuno-epidemiological models for both countries account for both the duration of natural and vaccinal immunity and the residual decrease in host susceptibility to infection (relative to immunologically naive individuals) after full natural or vaccinal immunity has waned; these models are described in detail in (*5, 6*), where the more detailed structure (*6*) accounts for immunity after one or two vaccine doses. In the first “decoupled” framework (top panel of Figure 1), we assume that the epidemiological dynamics of both countries are entirely independent with the exception of their respective vaccination rates, and we also compute a measure for the global potential for viral evolution of immune escape (*6*). In the second “coupled” framework (bottom panel of Figure 1), we allow for immigration of infected individuals between the countries at rate *η* (*17*). Additionally, we approximate the stochastic occurrence of potential transmission increases (PTIs) in each country: briefly, if the ‘potential net viral adaptation rate’ (see Supplementary Materials, Figure 5, and (*6*)) exceeds a threshold, then there is a non-zero probability that the transmission rate in both countries increases. This follows evidence of enhanced binding of the SARS-CoV-2 spike protein receptor binding domain (RBD) with the ACE2 receptor in more contagious SARS-CoV-2 variants, as well as potentially higher viral loads (*21*). Structural changes to the spike protein furin cleavage site (e.g. at site 681) may also contribute to increased viral transmissibility (*22*). In this way, this assumption represents a pessimistic scenario where the evolution of pathogen immune escape is inevitably accompanied by increases in transmission, although we also compare our results with the more optimistic scenario where transmission increases do not occur (note that we are not modeling the complexities of variant dynamics and evolution explicitly). In reality, transmission increases may plateau, and viral evolution may have more subtle effects on disease transmission including modulating the susceptibility of partially immune hosts. The full mathematical details for both frameworks are described in the Supplementary Materials.

**Figure 1:**
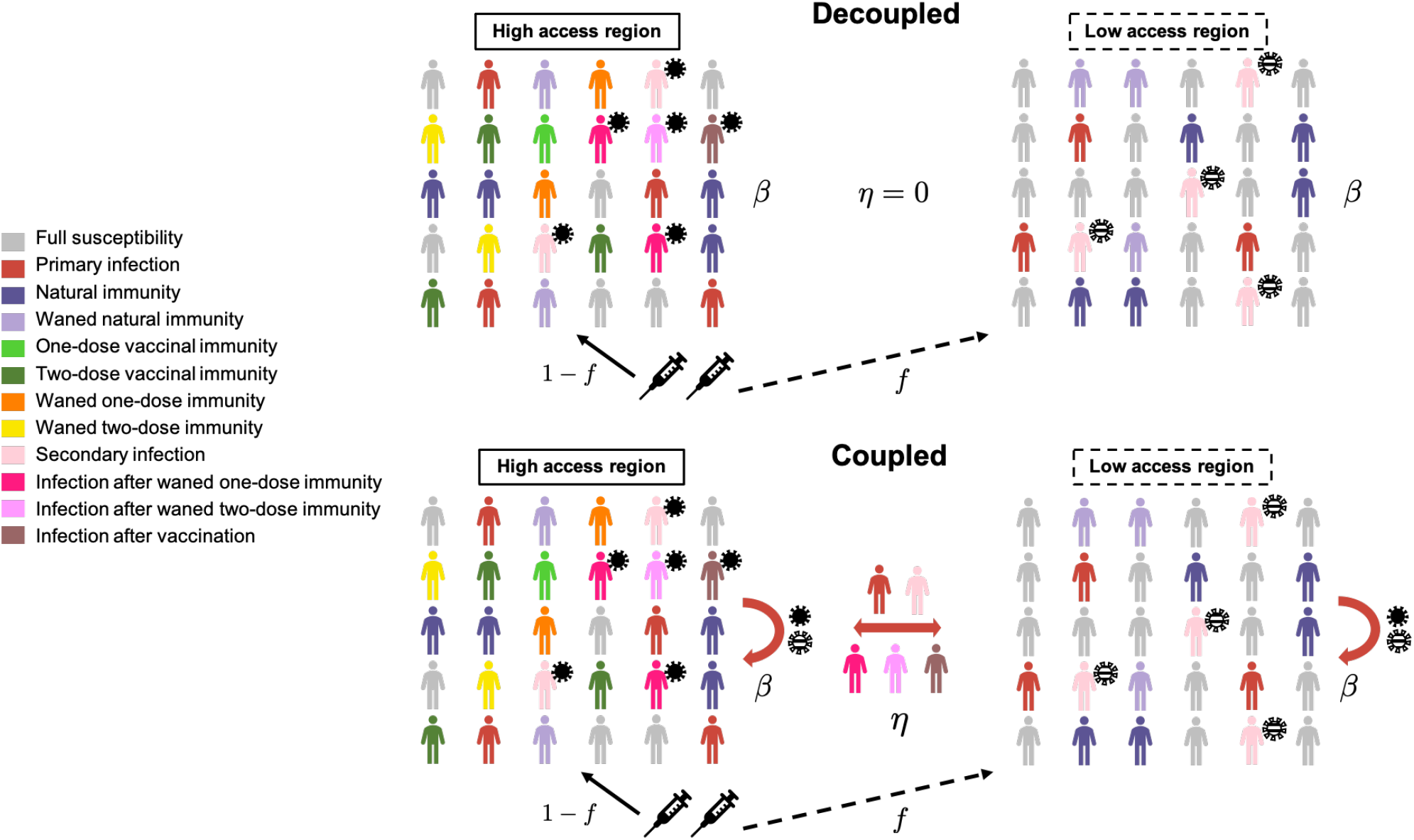
Schematic depicting the two-country model. The underlying immuno-epidemiological models for each country are based on (*5, 6*). Vaccines are allocated by the high access region (HAR) to the low access region (LAR). In the coupled framework, immigration of infected individuals between the countries is considered, and the national transmission rate depends on potential transmission increases (PTIs) in both countries, shown schematically as solid and striped virus particles in the HAR and LAR, respectively. In the decoupled framework, no immigration occurs, and the transmission rate is not influenced by PTIs. Full model details are provided in the Supplementary Materials.

We begin with the decoupled framework and the simpler underlying ‘one-dose’ vaccination model from (*5*) to compute the long-term equilibrium fraction of infections in both countries under a range of epidemiological and immunological scenarios. Then, with specific dosing regimes (*6*), we examine the short- and medium-term epidemiological dynamics and the global potential for evolution with the sharing of vaccines. Next, using the coupled framework, we compute national and combined case numbers in the medium term given different degrees of vaccine allocation from the HAR to the LAR for different immigration rates and average relative reproduction numbers. We do so for total as well as severe cases, with the expectation that the number of severe cases may be indicative of infections requiring hospitalization or the clinical burden of Covid-19, while the number of total cases reflects all infections regardless of severity. Finally, we compare the results of the coupled and decoupled frameworks for specific scenarios.

## Results and Discussion

### Decoupled Framework

The dynamics of prophylactic vaccine distribution strategies are well understood when infections lead to recovery and lifelong immune protection (SIR) (*14*). However, natural and vaccinal immunity to SARS-CoV-2 is likely not lifelong, yet complete re-susceptibility after the waning of immunity, as is assumed in susceptible-infected-recovered-susceptible (SIRS) frameworks, is also unlikely. Thus, we generalize across the strength and duration of immune responses with appropriate mathematical models (*5, 6*). We first ignore the complexities of dosing regimes and extend the model in (*5*) to consider vaccine sharing in the decoupled framework (top panel of Figure 1) under the assumption that a single immune category exists for vaccinated individuals, and that vaccinal immunity may wane at a rate distinct from natural immunity. Since we assume that the infection dynamics in both countries are only coupled through their respective vaccination rates, a unique equilibrium of total infections exists (either disease-free or endemic; see supplement and (*5*) for details). To examine the long-term epidemiological effects of vaccine nationalism, we compute the total fraction of infections at equilibrium as the proportion of vaccines shared between countries is varied. In other words, for a fixed global vaccination rate *ν*_tot_ (determined by the maximal rate of administration of the first dose *ν*_0,tot_ and the inter-dose period, see Methods) and for a fraction *f* of vaccines allocated from the HAR to the LAR, the vaccination rates in the HAR and LAR are (1 *− f*)*ν*_tot_ and *fν*_tot_, respectively. We examine four immunity scenarios that range from very poor to robust natural and vaccinal immune responses (Figure 2).

**Figure 2:**
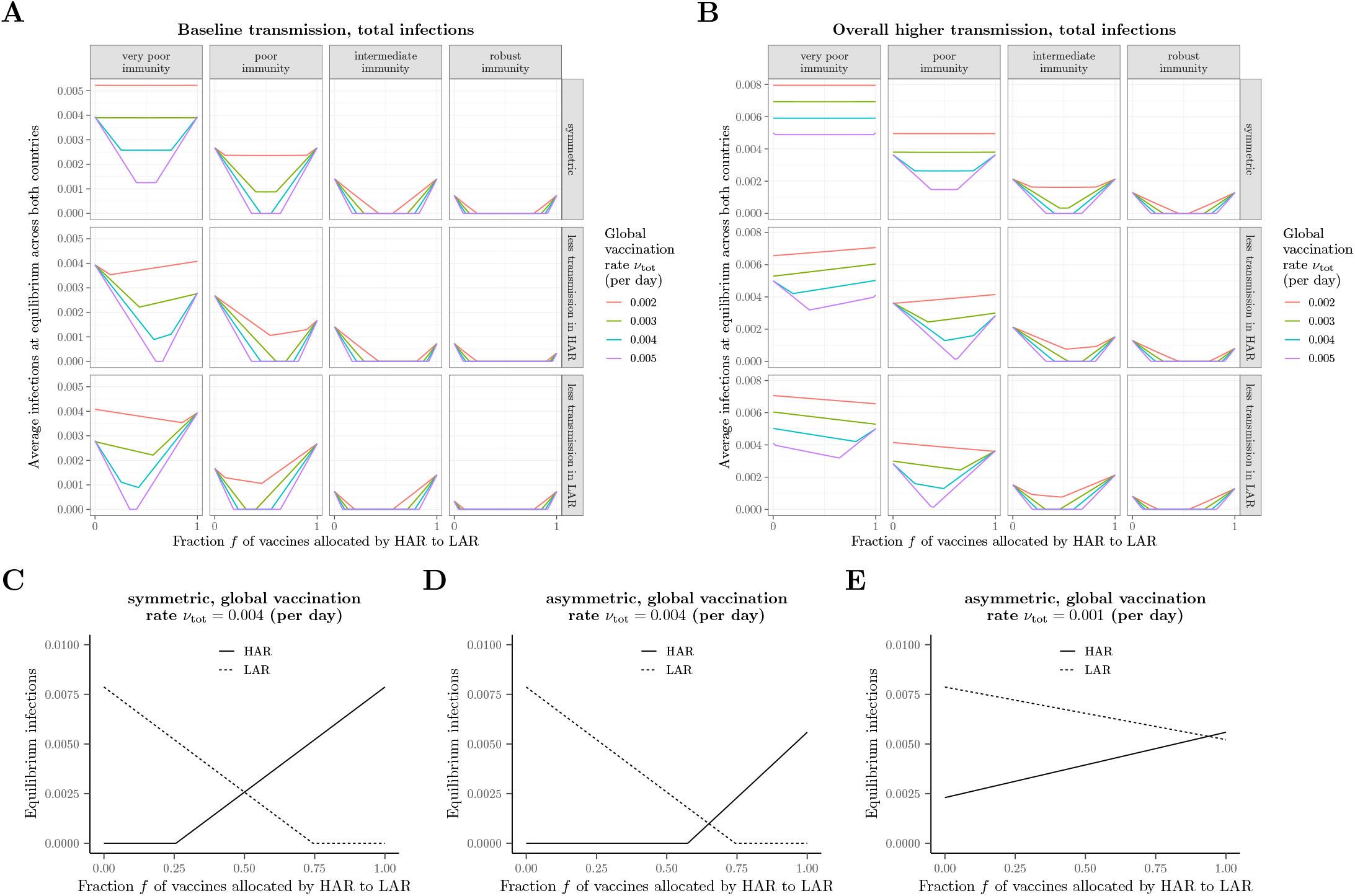
Long-term equilibrium of the average fraction of infections. In all panels, immunity scenarios are as follows: *very poor immunity*,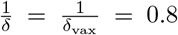 years, *ϵ* = 0.8; *poor immunity*,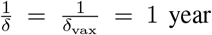 *ϵ=* 0.7; *intermediate immunity*,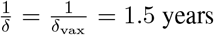, *ϵ* = 0.6; *robust immunity*,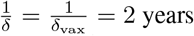, *ϵ* = 0.5. In the scenario with asymmetrical transmission rates between the two countries, the transmission rate in the country with lower transmission is taken to be 80% of the value in the symmetric case. In the scenarios with overall higher transmission rates (panel *B*), this same asymmetric assumption is made in addition to the baseline symmetric transmission rate being elevated by 30% relative to the value in panels (*A*). (*C*)–(*E*) Illustrations of the equilibrium fraction of infections in each country with the very poor immunity scenario, for: (*C*) symmetric transmission with *ν*_tot_ = 0.004; (*D*) asymmetric transmission (lower in HAR) with *ν*_tot_ = 0.004; (*E*) asymmetric transmission lower in HAR) with *ν*_tot_ = 0.001; with all other parameters as in panel *A*. In all panels, the baseline transmission rate is 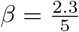.

When the characteristics of both countries are the same, sharing vaccines always decreases or maintains the total fraction of infections at the long-term equilibrium (see Keeling (*14*) for the SIR extreme with a focus on two differently-sized populations). The intuition for this result is apparent from examining the underlying values for each country (Figure 2C). The total fraction of infections are minimized whenever one of the countries does not vaccinate beyond the rate needed for herd immunity. Additionally, sharing does not have an appreciable impact on the total fraction of infections at equilibrium when vaccination rates are too low (Figure 2A, top panel), or overall transmission rates are more elevated and host immune responses are poorer (Figure 2B, top panel). Because of nonpharmaceutical interventions (NPIs) or intrinsic factors (e.g. population density (*23*) or vulnerabilities (*24*)), transmission rates in the two countries may be asymmetric. If there is less disease transmission in the HAR (modelled as a reduction in the transmission rate), then the ‘optimal’ fraction of vaccines shared to minimize the combined equilibrium fraction of infections crucially depends on the magnitude of the vaccination rate. If vaccine supplies are low and immune responses very poor, then sharing only a very small fraction of the vaccine supply is epidemiologically beneficial in terms of decreasing the overall burden. For stronger immune responses, augmenting vaccine sharing rates becomes increasingly beneficial from an epidemiological perspective, as the protective effects of the vaccine are maintained for longer within the population (compare the columns of the middle row of Figure 2A). Similarly, as vaccine supplies increase (compare the coloured curves in the middle row of Figure 2A), the minimum value of infections occurs for increasingly large values of *f*, or fractions of vaccines shared. Eventually, when global vaccination rates are high, even for very poor host immune responses, this minimum is attained when more than half of the vaccine supply is allocated to the LAR (leftmost panel of middle row of Figure 2A). By symmetry, the opposite occurs if there is less transmission in the LAR. These trends are further magnified if overall transmission rates are increased (Figure 2B). To further emphasize these effects, we present the long-term equilibrium of each country under representative scenarios in Figures 2C–2E. In particular, the comparison between Figures 2D and 2E illustrates the importance, and indirect benefit, of increasing vaccine supply. The relative sizes of the HAR and LAR populations can have important consequences for the fraction of vaccine allocation that minimizes the weighted fraction of infections (Fig. S1, S2). Overall, these results highlight the importance of continued NPIs that decrease transmission, such as rapid-testing and physical distancing, in conjunction with ramping up vaccination and sharing vaccine supplies equitably to decrease overall burden.

To consider the near- and medium-term dynamic epidemiological effects of vaccine sharing, in Figure 3 we explore the landscapes of immunity and infections across multiple scenarios for otherwise-symmetric countries (i.e. population size and seasonal transmission rates). In all scenarios, vaccine supply is assumed to be limited initially in the HAR (modeled as a one dose policy, a lower maximal rate of administration of the first dose *ν*_0,tot_, and no sharing (*f* = 0)) and then is assumed to increase. In conjunction with an increase in *ν*_0,tot_, we allow for a transition to the recommended two-dose strategy (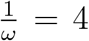, right two columns of Figure 3), and/or the initiation of equal sharing (*f* = 0.5) with the LAR (which is assumed to distribute vaccines using the same strategy as the HAR) may be initiated (second and fourth columns of Figure 3).

**Figure 3:**
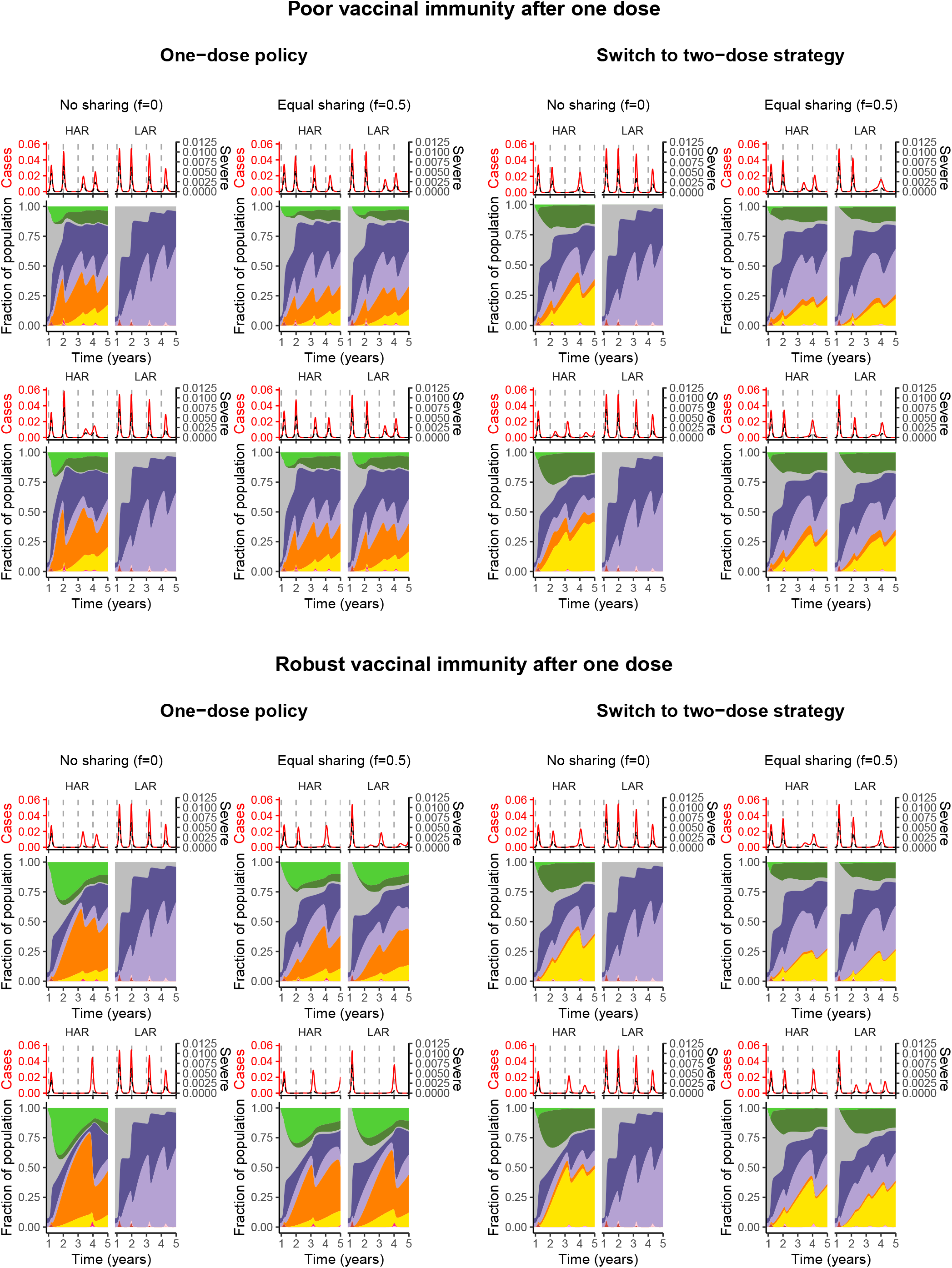
Immune landscapes and infections in both countries under a range of vaccination strategies and assumptions related to robustness of immune responses. Note that the color scheme is as in Figure 1. In all panels, vaccination begins after week 48.*Poor vaccinal immunity after one dose* is represented by 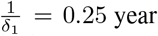 and *ϵ*_1_ = 0.9, whereas *robust vaccinal immunity after one dose* means 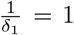 and *ϵ*_1_ = 0.7. All other parameters values including the procedure for the calculation of severe cases are described in Supplementary Materials. In both the top and bottom panels: the top row depicts a switch from a maximum first-dose administration rate of 1% to 3% after week 60, whereas it is 1% to 5% for the bottom row (and concurrent with sharing, if it occurs).

Intuitively, if one-dose immunity is robust (bottom panel of Figure 3), then transitioning to a two-dose strategy leads to fewer individuals with robust vaccinal immunity, in turn giving rise to substantial increases in infections in both in the short- and medium-term (compare the corresponding scenarios of the bottom panel of Figure 3 and see also (*6*)). In such a situation, ‘one-dose’ strategies (*i*.*e*. either the first dose of a 2-dose vaccine or the unique dose of a 1-dose vaccine) with equal sharing between countries suppress overall burden. On the other hand, if one-dose immunity is poor, switching to the recommended two-dose regimen prevents the accumulation of individuals with waned one-dose immunity and thus potentially larger infections peaks in the longer term (top panel, Figure 3). If poor one-dose immunity nevertheless reduces severity of infection after waning (unlike our pessimistic assumption), then the predicted clinical burden of severe cases would likely be lower. Finally, if one-dose immunity is poor and a one-dose policy is pursued, the first infection peak after ramping up vaccination in the HAR may be higher without sharing (top left panel of Figure 3). This counter-intuitive finding arises due to the large accumulation of individuals with waned one-dose immunity who experience infection. This highlights the important role for population-level susceptibility (modulated by natural and vaccinal immune responses) and its dynamical interplay with transmission in determining the timing and burden of infections. The effects of different NPI scenarios, transmission patterns, and vaccination rates in either the HAR or LAR can be further explored with the online application (https://grenfelllab.shinyapps.io/vaccine-nationalism/).

The accumulation of individuals with various immunity phenotypes (i.e. waned one-dose immunity or immunity following natural infection) may also lead to different evolutionary outcomes depending on the vaccine sharing scheme pursued. Current evidence suggests that adaptive immune responses following natural infection with SARS-CoV-2 are fairly robust and long-lasting (*25, 26*), although this may be less certain in the context of subsequent infection with variant strains (*27, 28*). Encouragingly, studies indicate that previously-infected hosts are largely protected (clinically and against breakthrough infections) against emerging variants after a single vaccine dose (*29*). Thus far, the duration and strength of this protection towards existing strains and potential emerging variants remain unknown. In Figure S3, we use the evolutionary framework from (*6*) to project the potential net viral adaptation rate (see Methods and online application for additional scenarios). Overall, we find that uncertainties in evolutionary outcomes dominate our projections, echoing previous findings (*6*) (Figure S3, and online application). However, if the evolutionary potential for immune escape is highest among infections in hosts with natural immunity, then sharing vaccines always decreases global evolutionary potential (Figure S3) in the decoupled framework. Overall, when immunity after a single vaccine dose is robust, natural and vaccine-derived immunity will limit evolutionary advances relative to the scenario with poor single dose immunity (compare the top and bottom panels of Figure S3). However, if immunity is partial or waning, ongoing transmission might accelerate adaption, supporting the need for continued monitoring of variants and their interaction with natural and vaccine-derived immunity.

Another intuitive result is that, in the LAR, sharing vaccines leads to increases in population immunity (for any dosing regime) and thus a decrease in infections and burden in the short term, even with poor one-dose immunity. In general, in the HAR, sharing decreases population immunity and increases infections in the short term. However, these changes are minimal and likely acceptable given the combined decrease in infections, illustrating the long term benefits of vaccine sharing.

### Coupled Framework

So far, we have assumed that the countries have decoupled disease dynamics. This simplification for tractability ignores infection importation as well as the possible emergence of variants from regions with more persistent infections. The issues that arise from the global circulation of SARS-CoV-2, particularly the variants of concern, are of considerable public health importance. Thus, we next explore these effects using the coupled framework presented in the bottom panel of Figure 1.

In Figure 4, we plot the cumulative number of total and severe cases (see Supplementary Materials for details) assuming equal population sizes in both countries from the time of vaccine introduction until 5 years after the pandemic onset in the HAR, LAR, and combined, as well as the projected number of PTIs to have occurred in both regions by the end of the 5 year period. We do so for various vaccine allocation fractions between the HAR and LAR, as well as a range of immigration rates assuming symmetric transmission rates (Figures 4A and 4C) and relative mean reproduction numbers assuming constant immigration rates (Figures 4B and 4D, see Methods). In Figures 4A and 4B we assume that infection following waned natural immunity contributes the most to viral adaptation, while in Figures 4C and 4D we assume that infection following waned vaccinal immunity, and one-dose immunity in particular, contributes the most.

**Figure 4:**
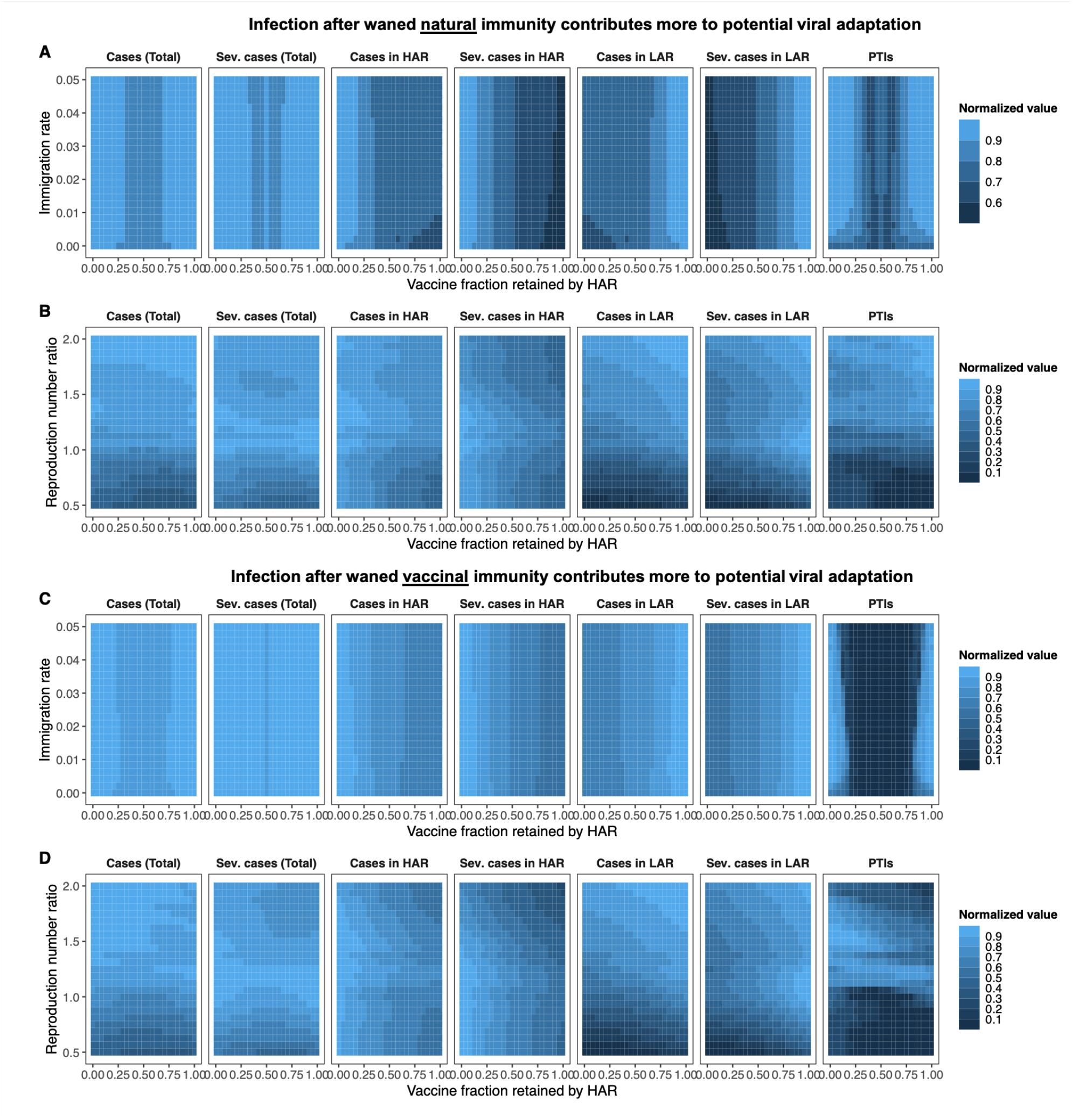
Heat maps depicting total and severe cases from the time of vaccine onset (*t*_vax_ = 48 weeks) through the end of the 5 year period for both countries (leftmost two columns), the HAR (third and fourth columns from the left), the LAR (fifth and sixth columns from the left), as well as the combined number of PTIs to have occurred in both countries at the end of 5 years (rightmost column). Each grid-point denotes the mean value of 100 simulations. The population of both countries is taken to be the same. Each area plot is internally normalized, such that the largest value in each plot is 1. The *x*-axis indicates the fraction of vaccines retained by the HAR (i.e. 1 − *f*); thus the far right of a plot is the scenario where the HAR retains all vaccines (*f* = 0). In A and C, both countries have the same average transmission rate (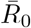, see Methods), and the immigration rate *η* is varied. In B and D, the immigration rate is fixed at *η* = 0.01, and the relative mean transmission rate in the LAR, i.e. 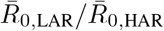, is varied between 0.5 and 2. The seasonality of the transmission rates in both countries and periods of NPI adoption are identical and as described in the Methods. In all simulations, we assume a two-dose strategy throughout, i.e.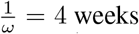, and take the maximal rate of administration of the first dose to be *ν*_0,tot_ = 2%. Assumed immunological parameters are 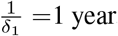, *ϵ*= 0.7, 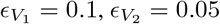, *ϵ*_2_ = 0.7,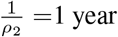, and the one-to two-dose immune response ratio is *x*_*e*_ = 0.8 (see Methods). In the top panel (A and B), we assume that infection after waned natural immunity contributes more to potential viral adaptation, and take *w*_*IS*_ = 0.8, *w*_*IS*1_ = 0.2*/x*_*e*_, and *w*_*IS*2_ = 0.2 (see Methods). In the bottom panel (C and D), we assume that infection after waned vaccinal immunity contributes more to potential viral adaptation, and take *w*_*IS*_ = 0.4, *w*_*IS*1_ = 0.8, and *w*_*IS*2_ = 0.8 × *x*_*e*_ (see Methods). Additional details related to the determination of severe cases are also provided in Supplementary Materials.

For equal population sizes and symmetric transmission rates, a weak dependence of total and severe cases as well as PTIs on the immigration rate *η* is observed, regardless of whether infection after waned natural or vaccinal immunity is assumed to contribute more to viral evolution (Figures 4A and4C). Additionally, equal vaccine sharing (i.e. *f* = 0.5) minimizes total PTIs and combined total cases in both scenarios. When natural infections contribute more to evolution (Figure 4A) and *η* is low, the HAR must retain an increasing share of the vaccines to minimize local total cases as the immigration rate increases, but this is done at the expense of more cases in the LAR and PTIs. Notably, for *f* ≈ 0, severe cases are minimized regardless of the immigration rate in the HAR and maximized in the LAR, which may have important clinical consequences. When infection following waned vaccinal immunity contributes more to viral evolution, this approach is no longer advantageous for the HAR, and the retained vaccine fraction sets the observed case numbers nearly independently of the assumed immigration rate. Further, large asymmetries in vaccine sharing (i.e. *f* = 0 or *f* = 1) result in much more marked relative numbers of PTIs in this scenario.

For the same total vaccine availability, the most realistic population asymmetry is that the LAR has a larger population; this corresponds to low and middle income countries with more fragile healthcare systems. Under this condition (Figure S4) and when infection after waned natural immunity contributes more to evolution, total cases in the LAR are relatively insensitive to the amount of vaccine allocated, except for very large *f*; however severe cases can be substantially reduced with vaccine sharing. Here, combined total cases and PTIs are minimized by minimizing cases in the HAR. When a greater number of total vaccines are available (i.e. a larger *ν*_0,tot_, Figure S5), total and severe cases in the more populous LAR decrease approximately monotonically with increasing vaccine allocation *f* and more equitable vaccine allocation once again minimizes combined total and severe cases. When infection after waned vaccinal immunity contributes more to evolution, the trends are more similar to those for symmetric population sizes, and more equitable vaccine sharing is favoured for both vaccination rates *ν*_0,tot_ given a larger population in the LAR (Figures S4 and S5).

When the LAR has a smaller population (Figure S6), a relatively weak dependence of total and severe case numbers and PTIs on *η* is still observed for both evolutionary scenarios, particularly for higher immigration rates. However, and particularly when infection after waned natural immunity contributes more to evolution, the minima in combined cases and PTIs are now observed for *f <* 0.5, i.e. when the HAR retains more than half of the available vaccines. Importantly, we note that additional booster doses may further change these landscapes of immunity, and consequently the projected burdens of total and severe cases.

The number of total and severe cases and PTIs show a greater sensitivity to the average reproduction number ratio 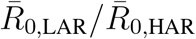 between the two countries for a fixed immigration rate. Intuitively, for equal population sizes (Figure 4) and when 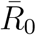 in the LAR country is lower, having the HAR retain more than half of the vaccines (*f <* 0.5) is a good strategy for minimizing total PTIs and cases, regardless of the evolutionary scenario. Indeed, the optimal vaccine allocation shifts closer and closer to equal sharing as the 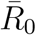 values of both countries approach each other, along with an increase in cases. These trends are similar when the LAR has a larger or smaller population than the HAR (Figures S4 and S6, respectively).

When 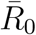 in the LAR is higher, trends are more complex. In general, regardless of the relative population size or evolutionary scenario, for a given 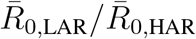, cases in the HAR decrease with increasing vaccine retention (smaller *f*), while cases in the LAR increase (Figures 4, S4, S5, and S6). Severe cases in each region in particular are strongly reduced by increased vaccine availability. The increase in cases in the LAR is increasingly large at higher 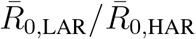. When infection after waned natural immunity contributes more to evolution and vaccine supply is sufficiently high to reduce case numbers in the LAR, PTIs are numerous when the HAR retains a large fraction of the vaccines (Figures 4B, S5B, and S6B). This is due to sustained elevated case numbers in unvaccinated individuals in the LAR. On the other hand, when infection following waned vaccinal immunity contributes more to evolution, then having the HAR (with lower transmission rates) retain a larger fraction of the vaccines minimizes PTIs for any relative population size, since the high 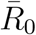 in the LAR would result in large subsequent peaks containing individuals whose vaccinal immunity has waned with sharing (Figures 4D, S4D, S5D, and S6D). However, this strategy also leads to highly elevated case numbers, including severe cases, in the LAR.

We note that assumptions of large 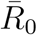 in the LAR also result in very large initial infection peaks, which increase community immunity in the medium-term. These initial waves are not reflected in the total case counts, however, since these values are summations from the time of vaccine initiation through the end of the 5 year period after the onset of the pandemic. Further, in Figure 5 we illustrate the temporal effect of the coupled framework on the infection dynamics in the LAR and HAR relative to the first model with no immigration or explicit effect of PTIs on the transmission rate. When moderate asymmetry in 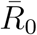 is assumed 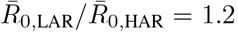, simulations using the decoupled framework suggest that a strategy where the HAR retains all vaccines would be highly beneficial for that country (top panel of Figure 5; no PTIs are projected to occur, and low case numbers are observed throughout). However, this occurs at the expense of PTIs and infection burden in the LAR, which are both substantially higher. With the more realistic coupled framework, immigration and increases in transmission illustrate that this strategy is far less beneficial to the HAR than the decoupled framework would suggest, as substantially higher case numbers and PTIs are predicted in this region. Although total cases in the HAR increase slightly when vaccines are equally distributed under the coupled framework (lower panel of Figure 5), substantial reductions in case numbers in the LAR result in fewer PTIs in that country, and total combined case numbers are also slightly lower. To untangle the effects of immigration and PTIs on dynamics in the coupled framework, we reproduce Figures 4 and 5 allowing for immigration only in Figures S7 and S8, respectively. In other words, these figures represent a more optimistic evolutionary scenario in which the occurrence of a PTI does not increase transmission rates in either the HAR or LAR. Overall, we show that vaccines play an important role in minimizing cases (particularly severe cases) as well as potential viral adaptation in both regions. We also emphasize that imperfect vaccinal and natural immunity and asymmetries in population size and transmission rates add many nuances to this picture.

**Figure 5:**
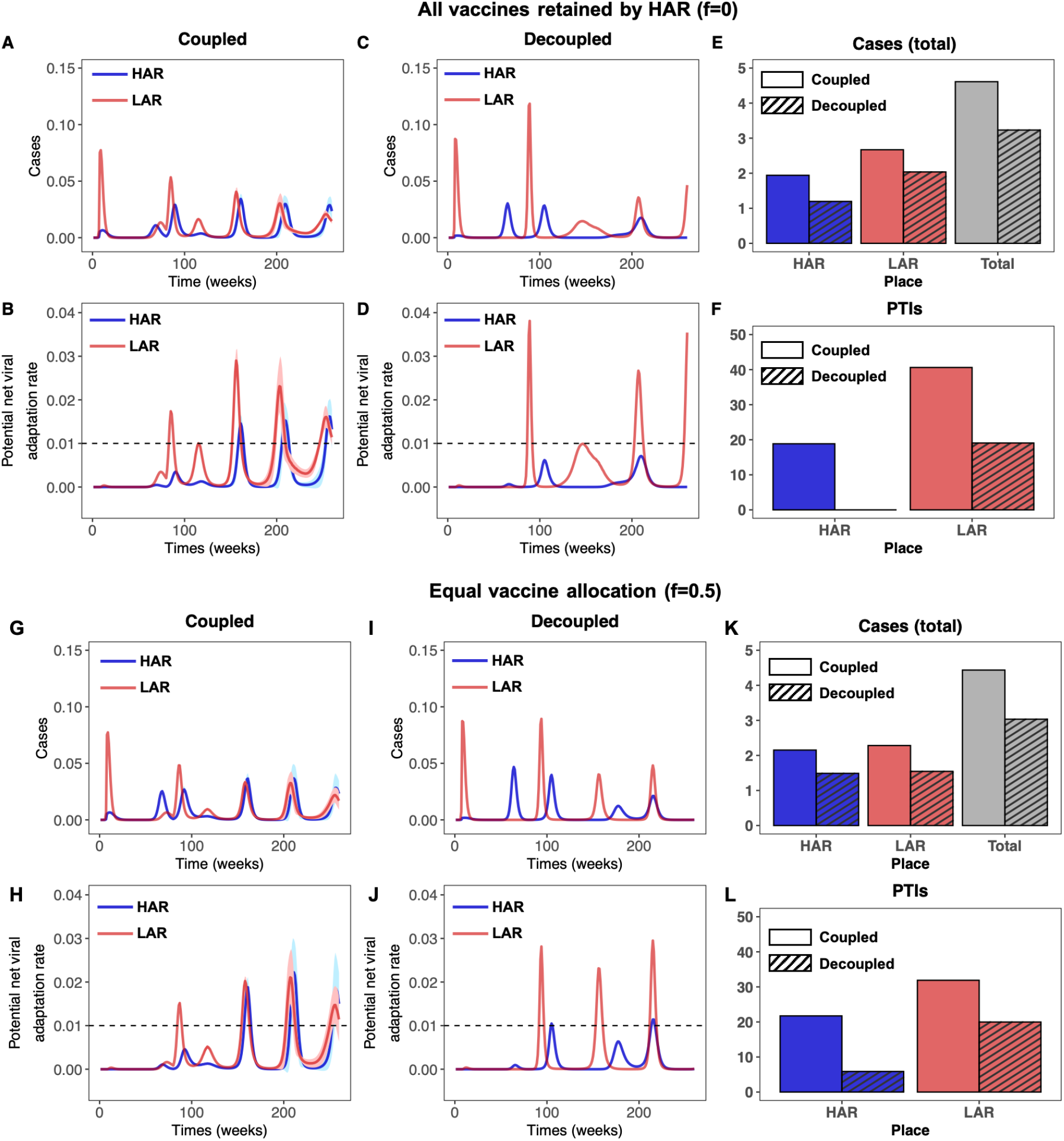
Time series of cases and potential net viral adaptation rates. Top row: Infections in the HAR (blue) and LAR (red) for the first 5 years after pandemic onset for the coupled (left) and decoupled (middle) frameworks. Each simulation is run 100 times, with the average indicated by the solid line and the standard deviation shown with the corresponding ribbon. The average number of cumulative cases over all simulations from the time of vaccine onset *t*_vax_ = 48 weeks through the end of the 5 year period are shown in the rightmost figure for the HAR, LAR, and both countries combined for the coupled (solid) and decoupled (dashed) frameworks. Bottom row: Time series of the potential viral adaptation rate in both regions for the decoupled (left) and upled (right) frameworks. The colors, averages and standard deviations are as described above. The dashed horizontal line denotes *e*_cutoff_ = 0.01, the assumed threshold for the occurrence of a PTI (see Methods). The average number of PTIs at the end of the 5 year period are shown in the rightmost figure for the HAR and LAR for the coupled (solid) and decoupled (dashed) frameworks. The top panel (A-F) corresponds to the HAR retaining all vaccines (*f* = 0), while the bottom panel (G-L) corresponds to equal vaccine sharing (*f* = 0.5). In all simulations, we take 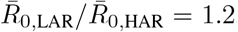, and assume that infection after waned natural immunity contributes primarily to evolution (i.e. *w*_*IS*_ = 0.8, *w*_*IS*1_ = 0.2*/x*_*e*_, and *w*_*IS*2_ = 0.2).All other parameters are identical to those in Figure 4.

### Caveats

A full list of caveats and future directions is presented in Supplementary Materials; we briefly summarize them below. First, building on prior work and in order to focus on qualitative features, we ignore heterogeneities within countries, such as due to age (*30*) or superspreading (*31*). Similarly, we have assumed simple scenarios for nonpharmaceutical interventions as in (*6*). More granular, well-parameterized epidemiological models with these complexities would lead to more accurate quantitative predictions. Furthermore, additional booster doses may be administered that could alter population-level immune landscapes, and including these in future models will be important for qualitative and quantitative predictions. Additionally, we omit vaccine hesitancy (*32*), though simple extensions of our previous models with hesitancy (*5, 6*) could examine the resulting interplay with vaccine nationalism. Furthermore, we have assumed the simplest evolutionary models, both for determining potential viral adaptation rates as well as for simulating potential increases in transmission rates. As more data become available, these should be refined (*33–36*), with possible directions including extending the model to explicitly track the transmission of different strains, and accounting for potential reductions in the strength of vaccinal immunity (i.e. the parameters 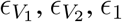, *ϵ*_1_, and *ϵ*_2_) due to the emergence of novel variants (*37*). Lastly, we assume that the seasonal transmission rates are similar in both countries, though in reality they could differ. The online interactive application (https://grenfelllab.shinyapps.io/vaccine-nationalism/) allows for an in-depth exploration of the effect of different climate-driven seasonal transmission rates as well as a broad range of assumptions related to NPIs and immuno-epidemiological parameters.

## Conclusion

Even as vaccine production increases, a number of countries are choosing to share little or no vaccines with countries that have very low vaccine availability. Vaccine nationalism, dosing regimes, and host immune responses have important interactive effects, and these will substantially shape epidemiological dynamics and evolutionary potential in the medium term. Additionally, unstable vaccine supply will also increase variability in the timing or availability of first and second doses.

Using extensions of our prior work (*5, 6*), we incorporated vaccine sharing scenarios in two countries whose infection dynamics are either otherwise independent or coupled through immigration of infectious individuals and evolution-driven increases in transmission rates. When country profiles are symmetric, we find that sharing vaccines with countries that have low availability decreases overall infections and may also mitigate potential antigenic evolution. Asymmetries in population size or transmission rates introduce additional complexities, which are particularly marked when natural and vaccinal immunity is weak. Nevertheless, our models indicate that the redistribution of vaccine surpluses is likely advantageous in terms of epidemiological and evolutionary outcomes in both countries and, by extension, globally. Ethical arguments also support this policy (*11, 12*). Persistent elevated disease transmission in countries with low vaccine availability also substantially undermines attempts at infection control via stockpiling in the country with high vaccine availability, which is not accounted for when disease transmission in both countries is assumed to be decoupled. Overall, our work highlights the importance of continued efforts in quantifying the robustness of immunity following vaccination. Furthermore, reevaluation of stockpiling policies as vaccine supplies increase is imperative, and ramping up global vaccination efforts is crucial.

## Data Availability

The manuscript does not include new data

## Supplementary Materials

See attached documents.

## Acknowledgements

This work was funded in part by Open Philanthropy, the Natural Sciences and Engineering Research Council of Canada through a Postgraduate-Doctoral Scholarship (CMSR), the Co-operative Institute for Modelling the Earth System (CIMES) (REB), the James S. McDonnell Foundation 21st Century Science Initiative Collaborative Award in Understanding Dynamic and Multi-scale Systems (CMSR, SAL), the C3.ai Digital Transformation Institute and Microsoft Corporation (SAL), Gift from Google, LLC (SAL), the National Science Foundation (CNS-2027908, CCF1917819) (SAL), the U.S. CDC (BTG), Flu Lab (BTG).

## Author contributions

CEW, CMSR, BTG designed the study. CEW and CMSR performed the simulations and analyses, and wrote the manuscript. SEM developed the Shiny application. All authors contributed to interpreting the results and editing the manuscript.

