## Supplementary Material for "Vaccine nationalism and the dynamics and control of SARS-CoV-2"

**Caveats and future directions** **8**

**Supplementary figures** **10**

**References** **17**

### Determination of seasonal reproduction numbers

In order to reflect observed seasonal variation in transmission rates for respiratory infections arising from related coronaviruses [1], influenza [1] and respiratory syncytial virus [2], we base seasonal reproduction numbers in this work on those in [3], which were calculated in [1] based on the climate of New York City. Other seasonal patterns can be explored using the interactive online application. In all simulations, we modify these values to force a mean value for the basic reproduction number of $\bar{R}_0 = \langle R_0(t) \rangle = 2.3$ by multiplying the climate-derived time series $R_{0,c}(t)$ by 2.3 and dividing by its average value, i.e.

$$R_0(t) = R_{0,c}(t) \frac{2.3}{\bar{R}_{0,c}}.$$

When transmission rates are assumed to be asymmetrical, the mean value of the $R_0(t)$ time series is adjusted by the desired relative ratio. For example, for $\bar{R}_{0,\mathrm{LAR}}/\bar{R}_{0,\mathrm{HAR}} = 1.2$, the reproduction number time series in the HAR is as above, but in the LAR it is

$$R_{0,\mathrm{LAR}}(t) = R_{0,c}(t) \frac{2.3}{\bar{R}_{0,c}} \frac{\bar{R}_{0,\mathrm{LAR}}}{\bar{R}_{0,\mathrm{HAR}}} = R_{0,c}(t) \frac{2.3}{\bar{R}_{0,c}} \times 1.2.$$

### Modeling of nonpharmaceutical interventions (NPIs)

In all simulations, we enforce periods of NPI adoption (arising from behaviours and policies such as lock downs, mask-wearing, and social distancing) in which the transmission rate is reduced from its seasonal value described in the previous section.

For the simulations using the decoupled framework (i.e. Figure 3), we used the same NPI scenarios as in [4]. For all simulations using the coupled framework (i.e. Figures 4, 5, S3, and S4), we assume that NPIs are adopted between weeks 8 and 44 following the pandemic onset resulting in the transmission rate being reduced to 40% of its seasonal value. Between weeks 45 and 79, we assume that the transmission rate is 60% of its seasonal value; higher than during the previous period due to either behavioural changes following the introduction of the vaccine or the emergence of more transmissible strains. Finally, we assume that NPIs are completely relaxed beyond week 80.

### Simple vaccination model

#### Model equations

For the equilibrium calculations, we use the model presented in [3] and assume vaccination is ongoing. For $i = A, B$, where for simplicity $A$ is the HAR and $B$ is the LAR, $I_{P,i}$ and $I_{S,i}$ denote the fraction of primary and secondary infections in country $i$, respectively. Similarly, for each country $i$, $S_{P,i}$ and $S_{S,i}$ denote the fraction of fully and partially susceptible individuals, respectively; $R_i$ is the fraction recovered from infection (fully immune); and $V_i$ is the fraction effectively vaccinated (also fully immune). Furthermore, $\beta_i$ and $\nu_i$ are the transmission rate and vaccination rate in country $i$, respectively. We assume that demographics and immune characteristics are the same in both countries: $\mu$ is the birth/death rate, $\gamma$ is the recovery rate, $\epsilon$ is the relative reduction in susceptibility following the waning of vaccinal or natural immunity, $\alpha$ is the relative infectiousness of secondary to primary infections, $\frac{1}{\delta}$ is the average duration of natural immunity, and $\frac{1}{\delta_{\text{vax}}}$ is the average duration of vaccinal immunity. The governing equations are thus

$$\frac{dS_{P,i}}{dt} = \mu - \beta_i S_{P,i}(I_{P,i} + \alpha I_{S,i}) - \mu S_{P,i} - \nu_i S_{P,i}, \tag{S1a}$$

$$\frac{dI_{P,i}}{dt} = \beta_i S_{P,i}(I_{P,i} + \alpha I_{S,i}) - (\gamma + \mu) I_{P,i}, \tag{S1b}$$

$$\frac{dR_i}{dt} = \gamma(I_{P,i} + I_S) - \delta R_i - \mu R_i, \tag{S1c}$$

$$\frac{dS_{S,i}}{dt} = \delta R_i + \delta_{\text{vax}} V_i - \epsilon \beta_i S_{S,i}(I_{P,i} + \alpha I_{S,i}) - \mu S_{S,i} - s_{\text{vax}} \nu_i S_{S,i}, \tag{S1d}$$

$$\frac{dI_{S,i}}{dt} = \epsilon \beta_i S_{S,i}(I_{P,i} + \alpha I_{S,i}) - (\gamma + \mu) I_{S,i}, \tag{S1e}$$

$$\frac{dV_i}{dt} = \nu_i(S_{P,i} + S_{S,i}) - (\delta_{\text{vax}} + \mu) V_i. \tag{S1f}$$

#### Equilibrium calculations

As computed in [3], we assume that $\alpha = 1$ (for all that follows), and the basic reproduction number [5, 6, 7] of country A or B is thus

$$\mathcal{R}_{0,i} = \frac{\beta_i}{\gamma + \mu}\left(\frac{\mu}{\nu_i + \mu} + \epsilon \frac{\nu_i}{\nu_i + \mu}\frac{\delta_{\text{vax}}}{\delta_{\text{vax}} + \nu_i + \mu}\right). \tag{S2}$$

As proved in [3], there is a unique equilibrium $\widehat{I}_{T,i} = \widehat{I}_{P,i} + \widehat{I}_{S,i}$ for each country (since they are decoupled), and so it follows that the model with simple vaccination (described by equations (S1a)-(S1f)) has a unique equilibrium. Within each country, if $\mathcal{R}_{0,i} < 1$, then this is the disease-free equilibrium. Otherwise, if $\mathcal{R}_{0,i} > 1$, the country's equilibrium has a positive fraction of total infections $\widehat{I}_{T,i}$. Note that the explicit formula for $\widehat{I}_{T,i}$ is given in [3]. For this analysis, since the two countries are only coupled through their vaccination rates, we can solve for the corresponding fractions of infections separately.

Both countries may not be the same size. Since the vaccination rate $\nu_{\text{tot}}$ is calibrated for country A, *i.e.* a fraction $\nu_{\text{tot}}$ of the population of country A is vaccinated per unit time, we let $\phi_{B,A}$ denote the relative size of country B to country A. Thus, assuming that a fraction $f$ of vaccines is shared from A to B, then the vaccination rates in each country are as follows:

$$\nu_A = (1 - f)\nu_{\text{tot}}, \tag{S3a}$$

$$\nu_B = \frac{1}{\phi_{B,A}} f \nu_{\text{tot}}. \tag{S3b}$$

Therefore, the weighted average of infections across both countries is

$$\widehat{I}_{T,\text{avg}} = \frac{1}{1 + \phi_{B,A}}\widehat{I}_{T,\text{tot}} = \frac{1}{1 + \phi_{B,A}}\left(\widehat{I}_{T,A} + \phi_{B,A}\widehat{I}_{T,B}\right). \tag{S4}$$

### Model with explicit dosing regimes

#### Model equations

Here, we use the model presented in [4], which is an extension of the model with simple vaccination described by equations (S1a)-(S1f). The additional classes of individuals in country $i$ are defined as follows: $V_{1,i}$ and $V_{2,i}$ denote those vaccinated with one or two doses, respectively; $S_{S_1,i}$ and $S_{S_2,i}$ denote those whose one- or two-dose vaccinal immunity has waned, respectively; $I_{S_1,i}$ and $I_{S_2,i}$ denote those infected after the waning of one- or two-dose vaccinal immunity, respectively (i.e. infected from the $S_{S_1,i}$ or $S_{S_2,i}$ class); and $I_{V,i}$ denotes those infected while in $V_{1,i}$ or $V_{2,i}$. There are also a number of additional parameters: letting $k = 1, 2$, $\epsilon_k$ is the relative reduction in susceptibility to infection following the waning of $k$-dose immunity; $\epsilon_{V_k}$ is the relative reduction in susceptibility while in $V_{k,i}$; $\frac{1}{\rho_k}$ is the average duration of $k$-dose immunity; $c$ is the fraction of individuals in $S_{S,i}$ for whom a single vaccine dose confers two-dose immunity; $x_k$ denotes the relative revaccination rate of individuals in $S_{S_k,i}$; $1 - p_k$ is the fraction of individuals in $S_{S_k,i}$ who obtain two-dose immunity after a single dose. The parameter $\nu_i$ now corresponds to the rate of administration of the first vaccine dose, and $\frac{1}{\omega_i}$ is the period between the first and second dose.

In addition to the model in [4], we also allow for coupling between the LAR and HAR through both the immigration rate $\eta$ as in [8] and potential increases in local transmission rates.

By defining the transmission-weighted total number of infections in each country $I_{T\alpha,i}$ as

$$I_{T\alpha,i} = I_{P,i} + \alpha I_{S,i} + \alpha_V I_{V,i} + \alpha_1 I_{S_1,i} + \alpha_2 I_{S_2,i},$$

the governing equations can be written as

$$\frac{dS_{P,i}}{dt} = \mu - \beta_i S_{P,i}[(1-\eta)I_{T\alpha,i} + \eta I_{T\alpha,j}\phi_{j,i}] - (s_{\text{vax}}\nu_i + \mu)S_{P,i}, \tag{S5a}$$

$$\frac{dI_{P,i}}{dt} = \beta_i S_{P,i}[(1-\eta)I_{T\alpha,i} + \eta I_{T\alpha,j}\phi_{j,i}] - (\gamma + \mu)I_{P,i}, \tag{S5b}$$

$$\frac{dR_i}{dt} = \gamma(I_{P,i} + I_{S,i} + I_{V,i} + I_{S_1,i} + I_{S_2,i}) - (\delta + \mu)R_i, \tag{S5c}$$

$$\frac{dS_{S,i}}{dt} = \delta R_i - \epsilon\beta_i S_{S,i}[(1-\eta)I_{T\alpha,i} + \eta I_{T\alpha,j}\phi_{j,i}] - (s_{\text{vax}}\nu_i + \mu)S_{S,i}, \tag{S5d}$$

$$\frac{dI_{S,i}}{dt} = \epsilon\beta_i S_{S,i}[(1-\eta)I_{T\alpha,i} + \eta I_{T\alpha,j}\phi_{j,i}] - (\gamma + \mu)I_{S,i}, \tag{S5e}$$

$$\begin{aligned}\frac{dV_{1,i}}{dt} = {} & s_{\text{vax}}\nu_i S_{P,i} + c s_{\text{vax}}\nu_i S_{S,i} + x_1 p_1 s_{\text{vax}}\nu_i S_{S_1,i} + x_2 p_2 s_{\text{vax}}\nu_i S_{S_2,i} - \\ & \epsilon_{V_1}\beta_i V_{1,i}[(1-\eta)I_{T\alpha,i} + \eta I_{T\alpha,j}\phi_{j,i}] - (\omega_i + \rho_1 + \mu)V_{1,i},\end{aligned} \tag{S5f}$$

$$\begin{aligned}\frac{dV_{2,i}}{dt} = {} & (1-c)s_{\text{vax}}\nu_i S_{S,i} + x_1(1-p_1)s_{\text{vax}}\nu_i S_{S_1,i} + x_2(1-p_2)s_{\text{vax}}\nu_i S_{S_2,i} \\ & + \omega_i V_{1,i} - \epsilon_{V_2}\beta_i V_{2,i}[(1-\eta)I_{T\alpha,i} + \eta I_{T\alpha,j}\phi_{j,i}] - (\rho_2 + \mu)V_{2,i},\end{aligned} \tag{S5g}$$

$$\frac{dI_{V,i}}{dt} = \beta_i(\epsilon_{V_1}V_{1,i} + \epsilon_{V_2}V_{2,i})[(1-\eta)I_{T\alpha,i} + \eta I_{T\alpha,j}\phi_{j,i}] - (\gamma + \mu)I_{V,i}, \tag{S5h}$$

$$\frac{dS_{S_1,i}}{dt} = \rho_1 V_{1,i} - \epsilon_1\beta_i S_{S_1,i}[(1-\eta)I_{T\alpha,i} + \eta I_{T\alpha,j}\phi_{j,i}] - (s_{\text{vax}}x_1\nu_i + \mu)S_{S_1,i}, \tag{S5i}$$

$$\frac{dS_{S_2,i}}{dt} = \rho_2 V_{2,i} - \epsilon_2\beta_i S_{S_2,i}[(1-\eta)I_{T\alpha,i} + \eta I_{T\alpha,j}\phi_{j,i}] - (s_{\text{vax}}x_2\nu_i + \mu)S_{S_2,i}, \tag{S5j}$$

$$\frac{dI_{S_1,i}}{dt} = \epsilon_1\beta_i S_{S_1,i}[(1-\eta)I_{T\alpha,i} + \eta I_{T\alpha,j}\phi_{j,i}] - (\gamma + \mu)I_{S_1,i}, \tag{S5k}$$

$$\frac{dI_{S_2,i}}{dt} = \epsilon_2\beta_i S_{S_2,i}[(1-\eta)I_{T\alpha,i} + \eta I_{T\alpha,j}\phi_{j,i}] - (\gamma + \mu)I_{S_2,i}. \tag{S5l}$$

For all simulations, we take $\mu = 0.02\text{y}^{-1}$ corresponding to a yearly crude birth rate of 20 per 1000 people. Additionally, we take the infectious period to be $1/\gamma = 5$ days, consistent with the modeling in [1, 3, 9] and the estimation of a serial interval of 5.1 days for Covid-19 in [10], and assume that $c = 0.5$. We take the relative transmissibility of infections to be $\alpha = \alpha_V = \alpha_1 = \alpha_2 = 1$, and therefore only modulate the relative susceptibility to disease $\epsilon$. For simplicity, we ignore re-vaccination and thus take $x_k = 0$. For the initial conditions of all simulations, we take $I_P = 1 \times 10^{-9}$ and assume the remainder of the population is in the fully susceptible class. The values of the remaining parameters used in the various simulations are specified throughout the main text.

#### Calculation of cumulative case numbers

The total number of cases at any time $t$ in a given country is calculated as

$$I_{T,i}(t) = I_{P,i}(t) + I_{S,i}(t) + I_{V,i}(t) + I_{S_1,i}(t) + I_{S_2,i}(t).$$

Cumulative case numbers between times $t_1$ and $t_2$ in a given country are then:

$$\gamma \sum_{t=t_1}^{t=t_2} I_{t,i}(t).$$

#### Linking vaccination rate to inter-dose period

As in [4], we consider an exponential relationship between the global rate of administration of the first vaccination dose $\nu_{\text{tot}}[\omega]$ and the inter-dose period $\frac{1}{\omega}$. We assume that the highest achievable rate $\nu_{0,\text{tot}}$ is attained when no second dose occurs (i.e. $\omega = 0$, an infinite inter-dose period), and that when the first and second doses are spaced by the clinically recommended inter-dose period $L_{\text{opt}}$ ($\omega_{\text{opt}} = \frac{1}{L_{\text{opt}}}$), the rate of administration of the first dose is one half of its maximum value. Thus,

$$\nu_{\text{tot}}[\omega] = 2^{-L_{\text{opt}}\omega}\nu_{0,\text{tot}}. \tag{S6}$$

#### Determination of potential net viral adaptation rate

The potential net viral adaptation rate $e_i$ in each country is calculated as in [4] as the sum of the sizes of the infection classes following waned immunity (i.e. $I_{S,i}$ after $S_{S,i}$, $I_{S_1,i}$ after $S_{S_1,i}$, and $I_{S_2,i}$ after $S_{S_2,i}$) weighted by the infection class-specific net viral adaptation rate. Specifically it is given by

$$e_i = w_{IS}I_{S,i} + w_{IS1}I_{S_1,i} + w_{IS2}I_{S_2,i}. \tag{S7}$$

#### Specification of one-dose vaccine efficacy from one- to two-dose immune response ratio

As in [4], the one- to two-dose immune response ratio $x_e$ sets the value of $\epsilon_1$ and $\rho_1$, the susceptibility to infection following waned one-dose vaccinal immunity and the duration of one-dose vaccinal immunity, respectively. Specifically, we take $\epsilon_1 = \epsilon_2 + (1 - x_e)(1 - \epsilon_2)$ such that the susceptibility to infection after a waned single dose interpolates linearly between the value after waned two doses ($\epsilon_2$) when the one and two dose immune responses are equally strong ($x_e = 1$) and unity (full susceptibility) when a single dose offers no immune protection ($x_e = 0$). Additionally, $\rho_1$ is given by $\rho_1 = \rho_2/x_e$.

#### Calculation of the number of severe cases

At any time $t$, we compute the number of severe cases in country $i$ as

$$I_{T,\text{sev},i}(t) = x_{\text{sev,p}}I_{P,i}(t) + x_{\text{sev,s}}I_{S,i}(t) + x_{\text{sev,V}}I_{V,i}(t) + x_{\text{sev,1}}I_{S_1,i}(t) + x_{\text{sev,2}}I_{S_2,i}(t).$$

In all applicable figures, we take $x_{\text{sev,p}} = 0.14$, $x_{\text{sev,s}} = 0.07$, $x_{\text{sev,V}} = 0.14$, and $x_{\text{sev,2}} = 0$. In Figure 3 of the main text, we take $x_{\text{sev,1}} = 0.14$ and $x_{\text{sev,1}} = 0$ for poor and robust one-dose vaccinal immunity, respectively. In Figure 4 of the main text and Figures S4-S7, we take $x_{\text{sev,1}} = x_{\text{sev,2}} + (1 - x_e)(x_{\text{sev,V}} - x_{\text{sev,2}})$ as in [4].

#### Details for coupling between the countries

##### Determination of immigration rate

The chosen immigration rate $\eta$ sets the degree of direct coupling between both countries via the importation of infected individuals. To account for the possibility of reductions in travel when NPIs are in place, at each time point we multiply the baseline value of $\eta$ by the largest value of the contemporary reductions in transmission rate in each country due to NPIs. For example, if at time $t$ the HAR has NPIs in place that reduce transmission to 40% of its seasonal value while the LAR has NPIs in place that reduce transmission to 60% of its seasonal value, $\eta$ is reduced to 40% of its baseline value. When the decoupled framework is considered, we set $\eta = 0$.

##### Estimation of occurrence of PTIs

At each time point, the potential net viral adaptation rate $e_i$ (defined previously [4]) is calculated in each country. We impose a threshold value of $e_{\text{cutoff}} = 0.01$, and if $e_i \geq e_{\text{cutoff}} = 0.01$, we allow for a PTI to occur with a 10% probability. Practically, we insure an integer number of PTIs by using the "Euler" integration method of the `R` ordinary differential equation (ODE) solver `rk`. We fix the initial step size at `hini`=0.1, and if a PTI does occur we set the time derivative of the number of PTIs to be 10, such that one PTI is accounted for in the specified time interval.

##### Linking the transmission rate to PTIs

At each time $t$ in each country, the baseline transmission rate $\beta_{i,0}(t)$ (determined as the product of the inverse of the infectious period $\gamma$, the local contemporary seasonal reproduction number $R_0(t)$, and any reductions due to the presence of NPIs) is assumed to be modulated by the total combined number of PTIs $N_{\text{tot}}(t)$ that have occurred, i.e. $N_{\text{tot}}(t) = N_A(t) + N_B(t)$. Specifically, we obtain the instantaneous transmission rate as

$$\beta_i(t) = \beta_{i,0}(t) \times 1.01^{N_{\text{tot}}(t)}.$$

When the decoupled framework is considered, we set $\beta_i(t) = \beta_{i,0}(t)$, and thus the occurrence of PTIs does not affect the transmission rate.

#### Caveats and future directions

In our work, we explore the simplest models of vaccine sharing, immunity, coupling, and evolution. There are a number of caveats, which we have summarized in the main text and expand on below. These are all important areas of future research.

- Our models make the simplest evolutionary assumptions. As more data are collected, more sophisticated models (e.g. [11, 12, 13]) should be formulated and incorporated into our framework. Specifically, we assume in this work that evolution results in secular increases in avidity to the ACE2 receptor leading to enhanced transmission following evidence in [14]. In reality, this effect may plateau, and the evolution of new variants may have more subtle effects on epidemiological parameters, including altering the effectiveness of vaccines [15]. As more data is accrued, the within-host evolution of SARS-CoV-2 and its connection to population-level disease transmission should be examined and carefully modelled (e.g. for influenza, see [16]).
- We also do not consider within-country heterogeneities, such as vaccine refusal [17], age [18], and superspreading [19]. In prior work, we have considered simple vaccine refusal extensions of our single-country models [3, 4], and similar extensions could be formulated to incorporate these factors.
- Additionally, we do not explicitly model immigration, and rather consider implicit contact between individuals in each country. Thus, incorporating explicit movement of individuals between regions (e.g. [20]) is an important area of future work that would increase the realism of the framework. Furthermore, we only consider two countries, but in reality the complex patterns of viral prevalence globally will be affected by vaccine nationalism. Thus, incorporating multiple countries into the model would be an important extension to further explore vaccine distribution policies.
- The NPI scenarios we consider are simple and fixed for certain durations. In reality, these are constantly changing and are highly dependent on human behaviour. Thus, incorporating human behaviour in our models (e.g. [21, 22, 23, 24, 25, 26]) is an important research direction.
- We also assume constant vaccination rates; however, as vaccine production increases, vaccination rates correspondingly increase. Thus, our model could be coupled with appropriate projections of time-dependent vaccination rates (themselves based on projected supply, e.g. as in [27]) to refine model predictions.
- For a number of reasons, transmission rates in different countries may have different patterns, e.g. due to climate [1], or inherent population-level vulnerabilities [28]. We examined some simple examples of this complexity in the main text, and further explorations are possible with the online interactive application. With proper calibrations to

various countries, future models should closely examine the ensuing epidemiological and evolutionary implications of transmission heterogeneities among countries.

- We have also assumed that infectious periods and immune durations are exponentially distributed. However, the distributions for both of these periods may have different shapes, and these could lead to complex epidemiological dynamics [29, 30]. Thus, investigating the resulting dynamics of our model with different distributions for infectious periods and durations of natural and vaccinal immunity would be important.

### Supplementary figures

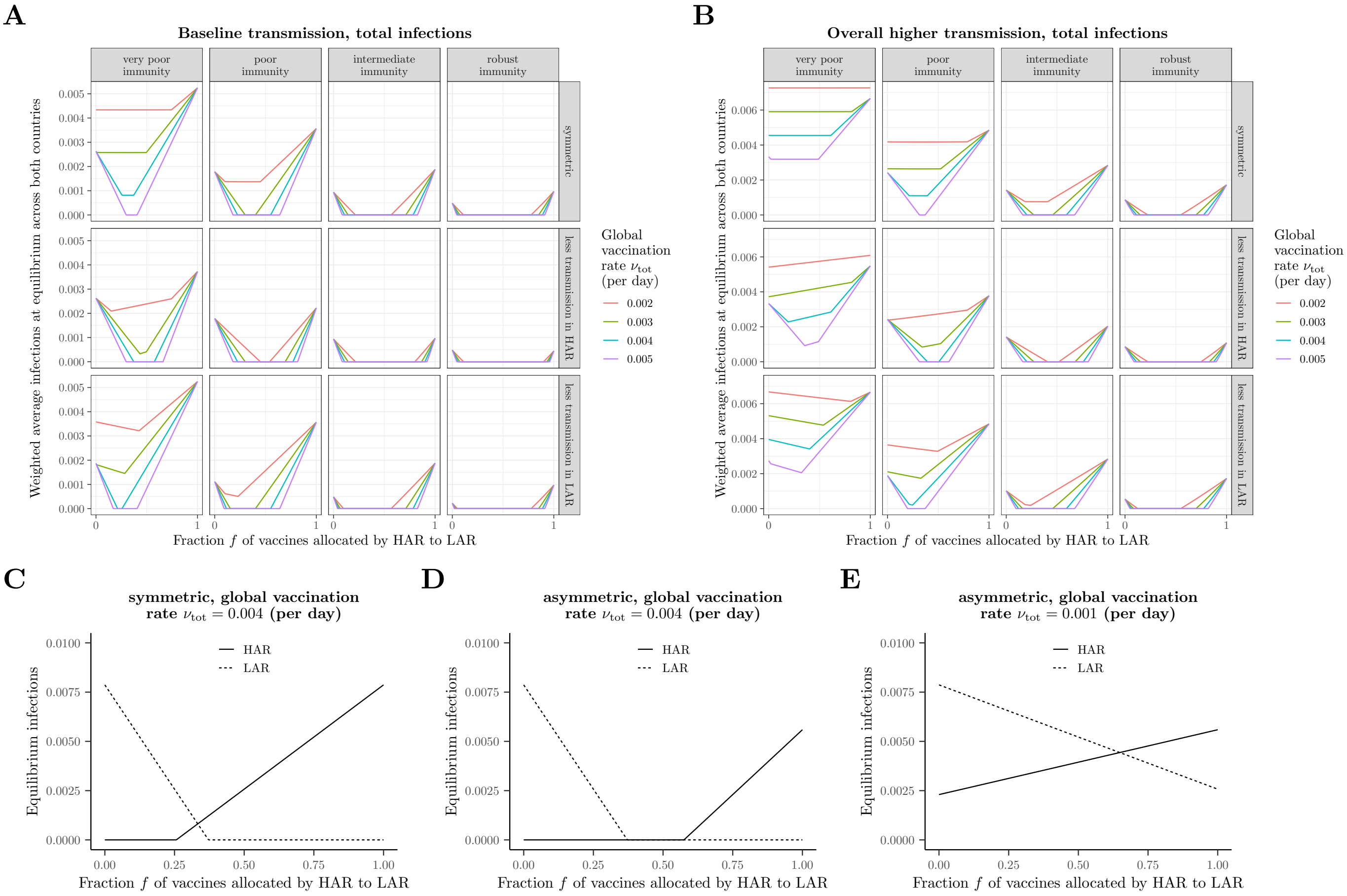

**Figure S1: Weighted average of combined equilibrium infections for a smaller LAR relative to HAR, i.e. $\phi_{B,A} = 0.5$ in the simpler model with vaccination. All other parameters are as in Figure 2, and scenarios (C)–(E) are identical to the corresponding scenarios presented in Figure 2C–2E.**

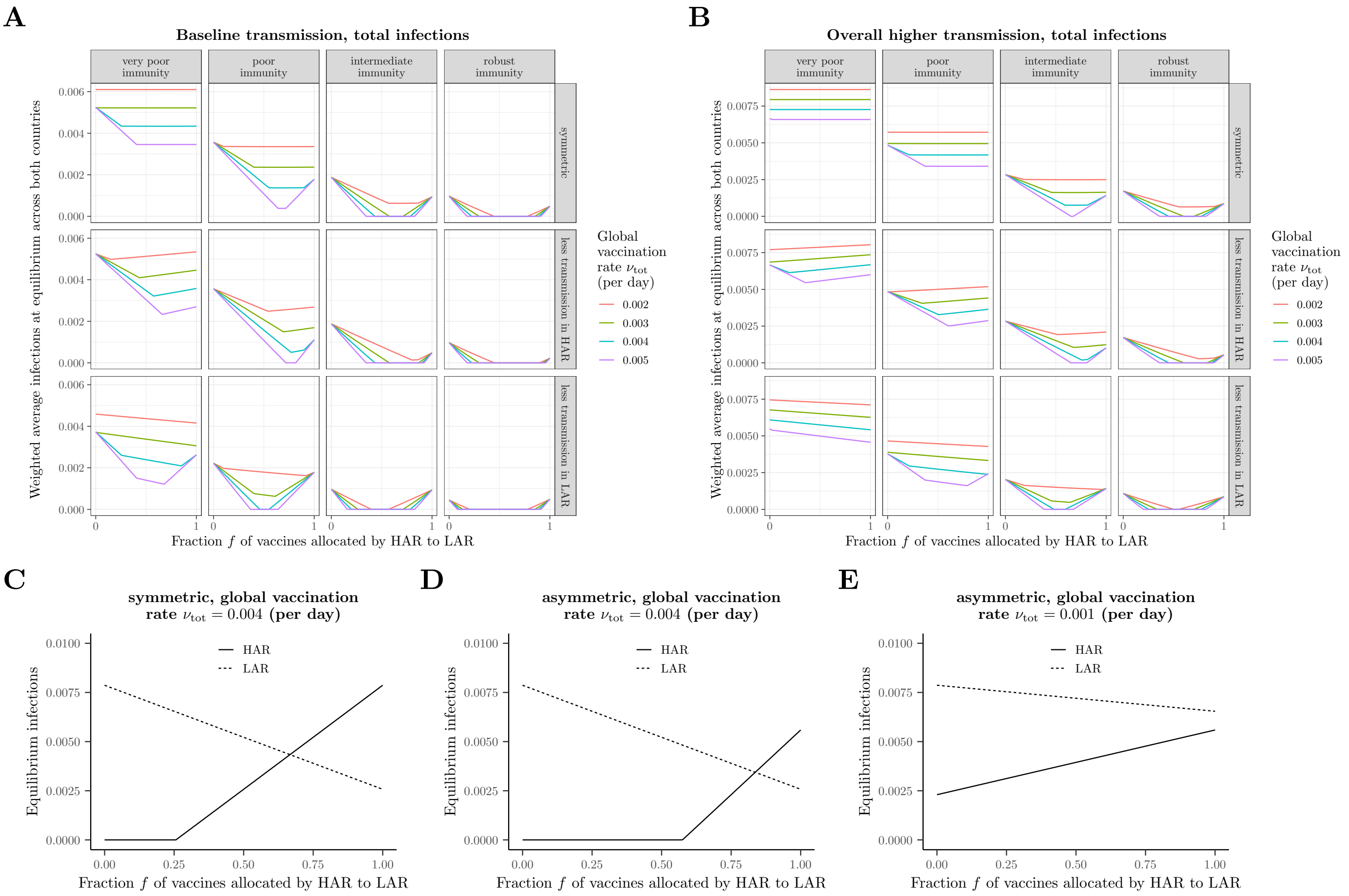

**Figure S2: Weighted average of combined equilibrium fraction of infections for a larger LAR relative to HAR, i.e. $\phi_{B,A} = 2$ in the simpler model with vaccination. All other parameters are the same as in Figure 2, and scenarios (C)–(E) are identical to the corresponding scenarios presented in Figure 2C–2E.**

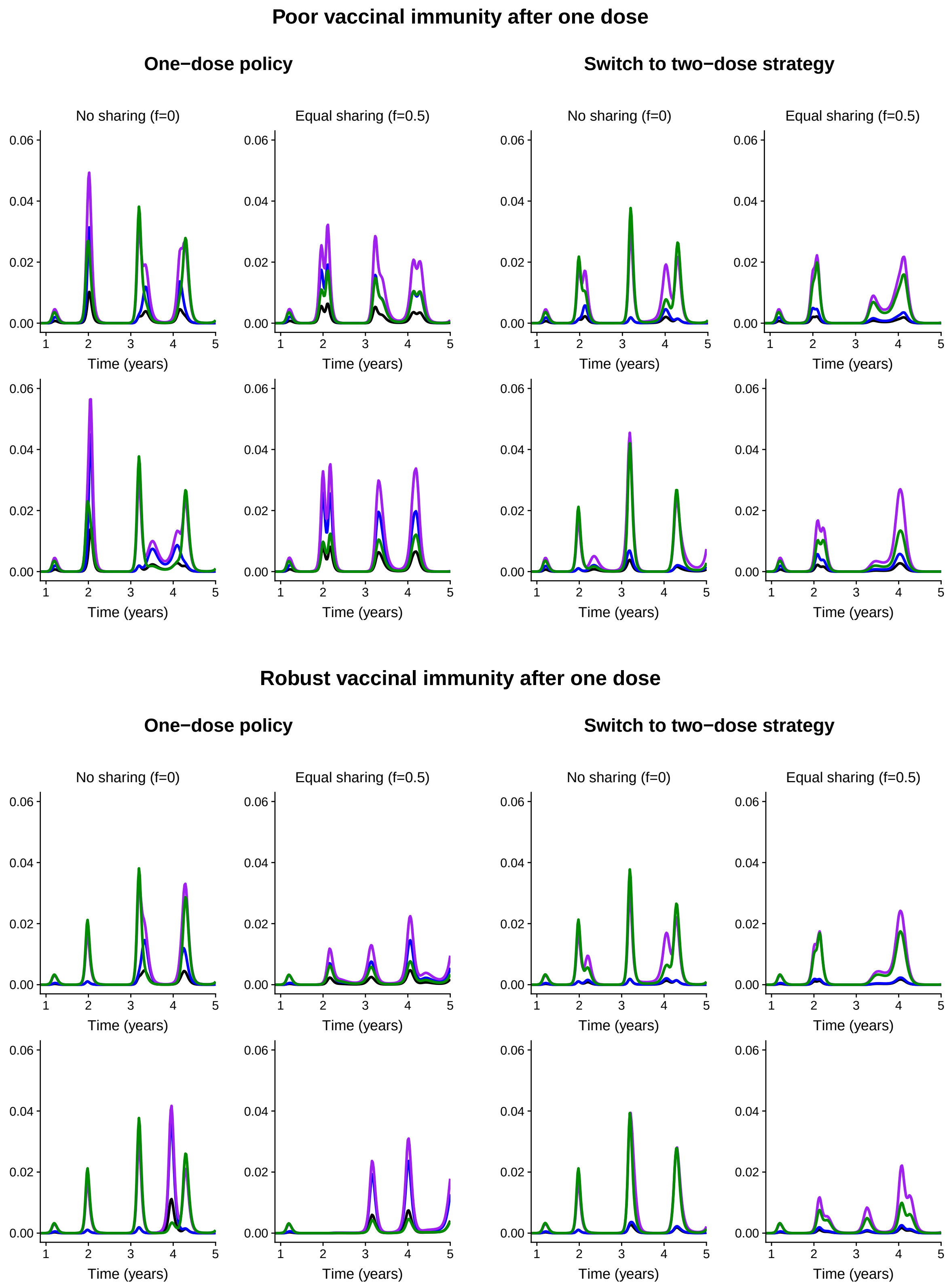

**Figure S3: Global evolutionary potential in the decoupled framework for the scenarios presented in Figure 3. The weights for each type of infection are (black) $w_{IS,I} = 0.05$, $w_{IS1,I} = 0.3$, $w_{IS2,I} = 0.05$; (blue) $w_{IS,I} = 0.05$, $w_{IS1,I} = 1$, $w_{IS2,I} = 0.05$; (purple) $w_{IS,I} = 0.8$, $w_{IS1,I} = 1$, $w_{IS2,I} = 0.8$; and (green) $w_{IS,I} = 1$, $w_{IS1,I} = 0.05$, $w_{IS2,I} = 0.05$.**

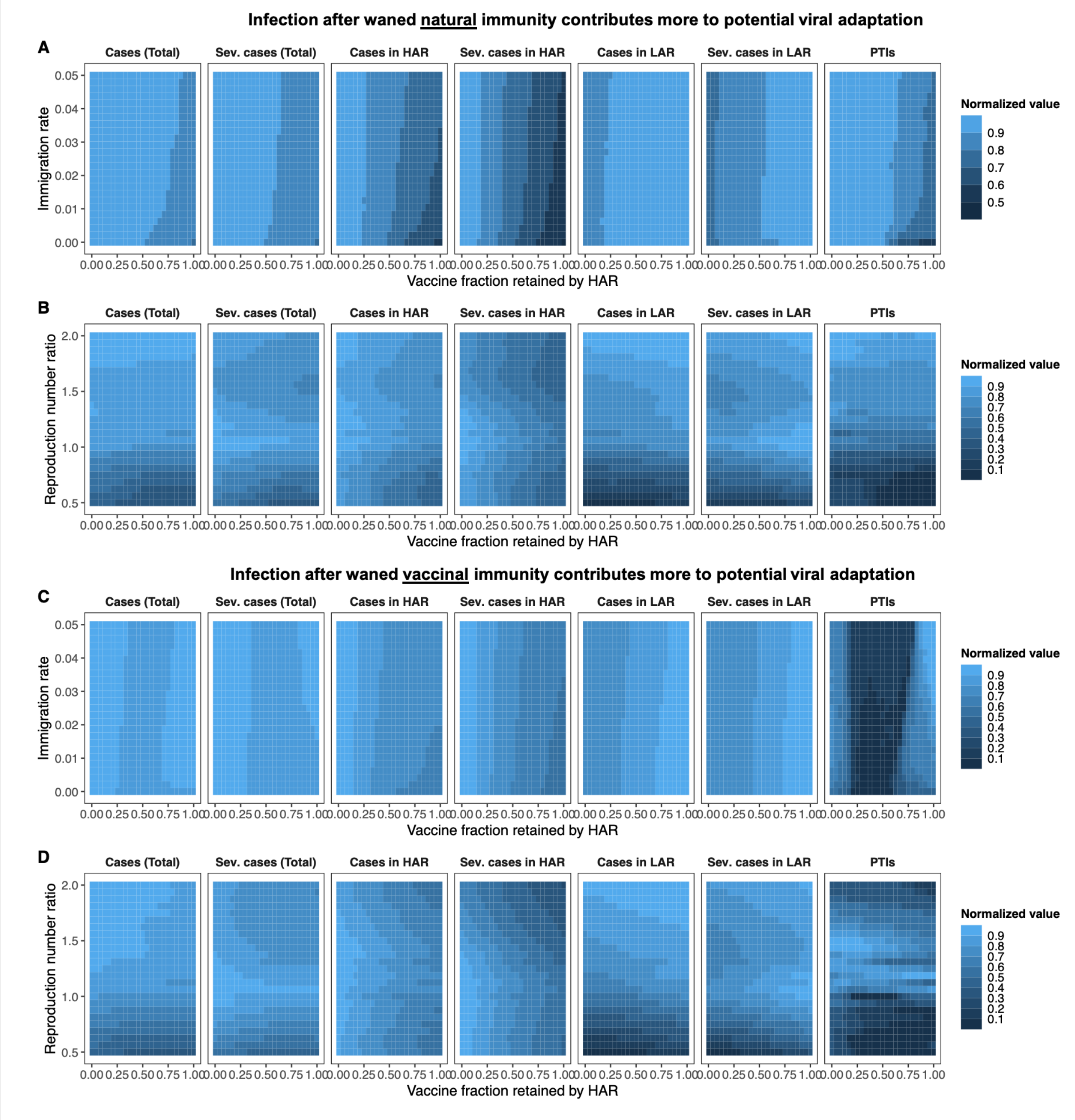

Figure S4: Heat maps depicting total and severe cases from the time of vaccine onset through the end of the 5 year period for both countries, the HAR, the LAR, as well as the combined number of PTIs to have occurred in both countries at the end of 5 years. All parameters are identical to Figure 4 of the main text, except the LAR is assumed to have a population double the size of the HAR, i.e. $\phi_{B,A} = 2$.

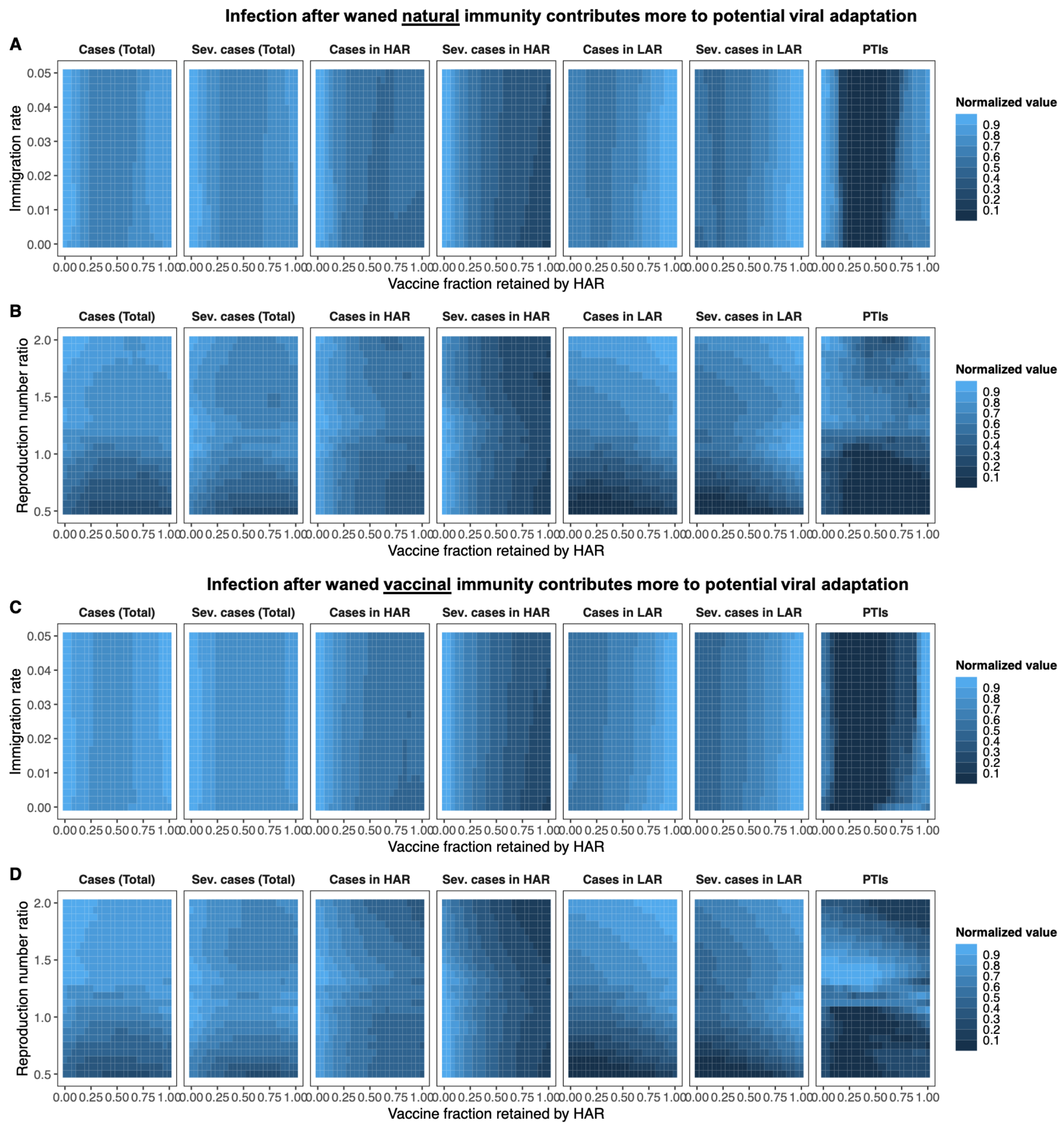

**Figure S5: Heat maps depicting total and severe cases from the time of vaccine onset through the end of the 5 year period for both countries, the HAR, the LAR, as well as the combined number of PTIs to have occurred in both countries at the end of 5 years. All parameters are identical to Figure 4 of the main text, except the LAR is assumed to have a population double the size of the HAR, i.e. $\phi_{B,A} = 2$, and vaccine availability is assumed to be greater: the maximal rate of administration of the first dose is set to $\nu_{0,\mathbf{tot}} = 5\%$ .**

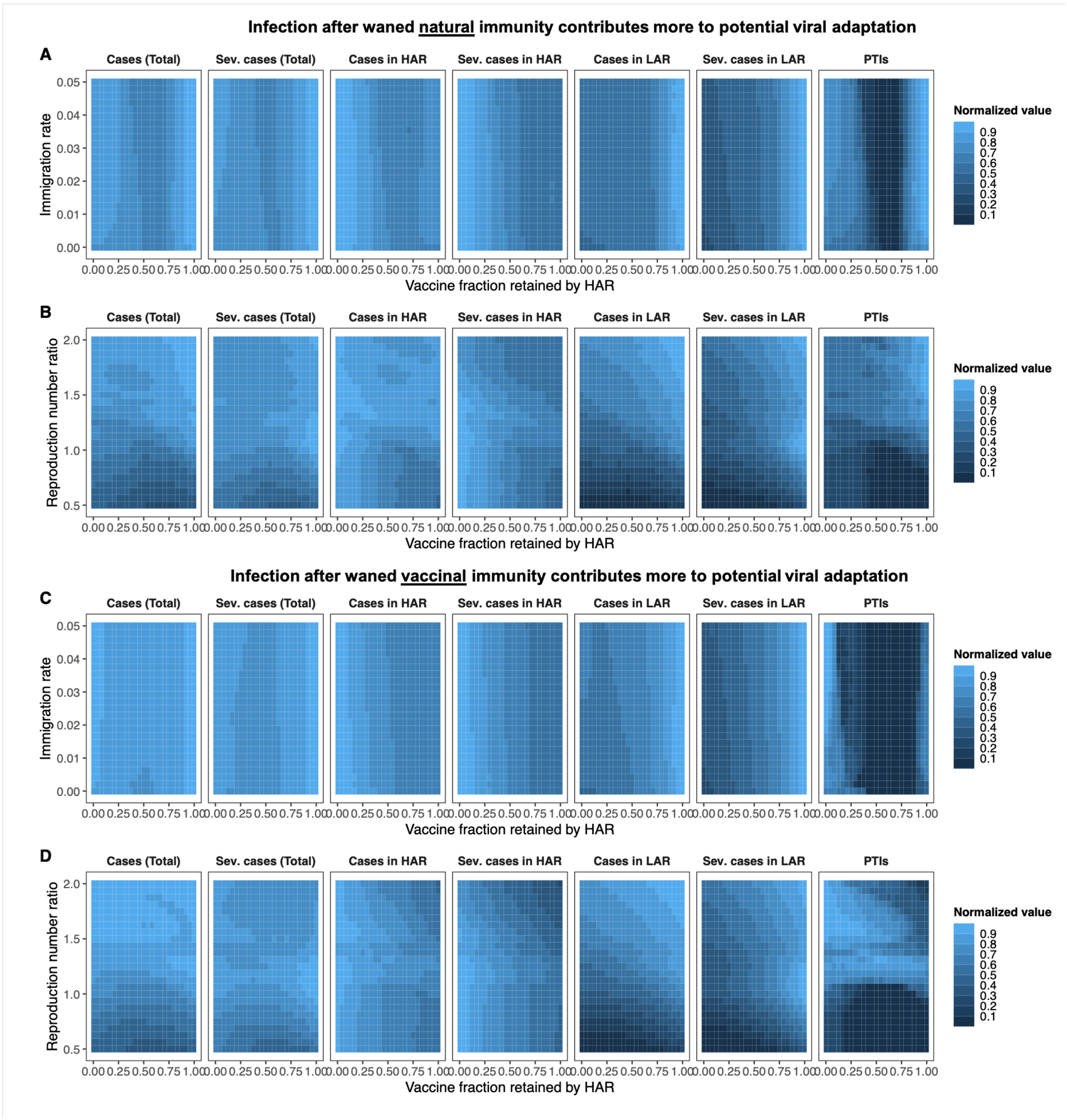

Figure S6: Heat maps depicting total and severe cases from the time of vaccine onset through the end of the 5 year period for both countries, the HAR, the LAR, as well as the combined number of PTIs to have occurred in both countries at the end of 5 years. All parameters are identical to Figure 4 of the main text, except the LAR is assumed to have a population one half of the size of the HAR, i.e. $\phi_{B,A} = 0.5$.

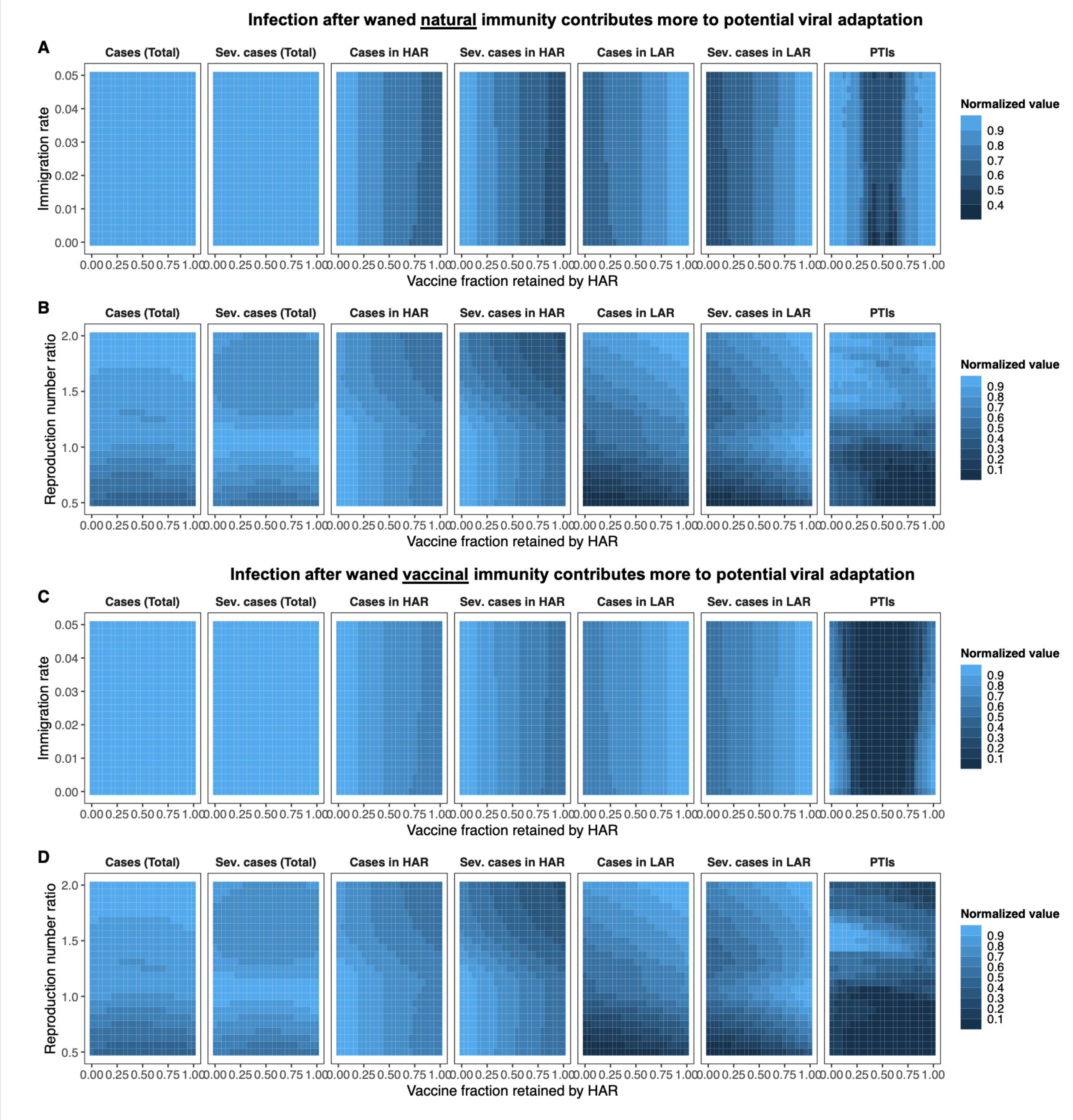

**Figure S7: Heat maps depicting total and severe cases from the time of vaccine onset through the end of the 5 year period for both countries, the HAR, the LAR, as well as the combined number of PTIs to have occurred in both countries at the end of 5 years. All parameters are identical to Figure 4 of the main text, except the occurrence of a PTI is assumed to not increase the transmission rate, i.e. $\beta_i(t) = \beta_{i,0}(t)$ in both the HAR and LAR for all times (see Methods).**

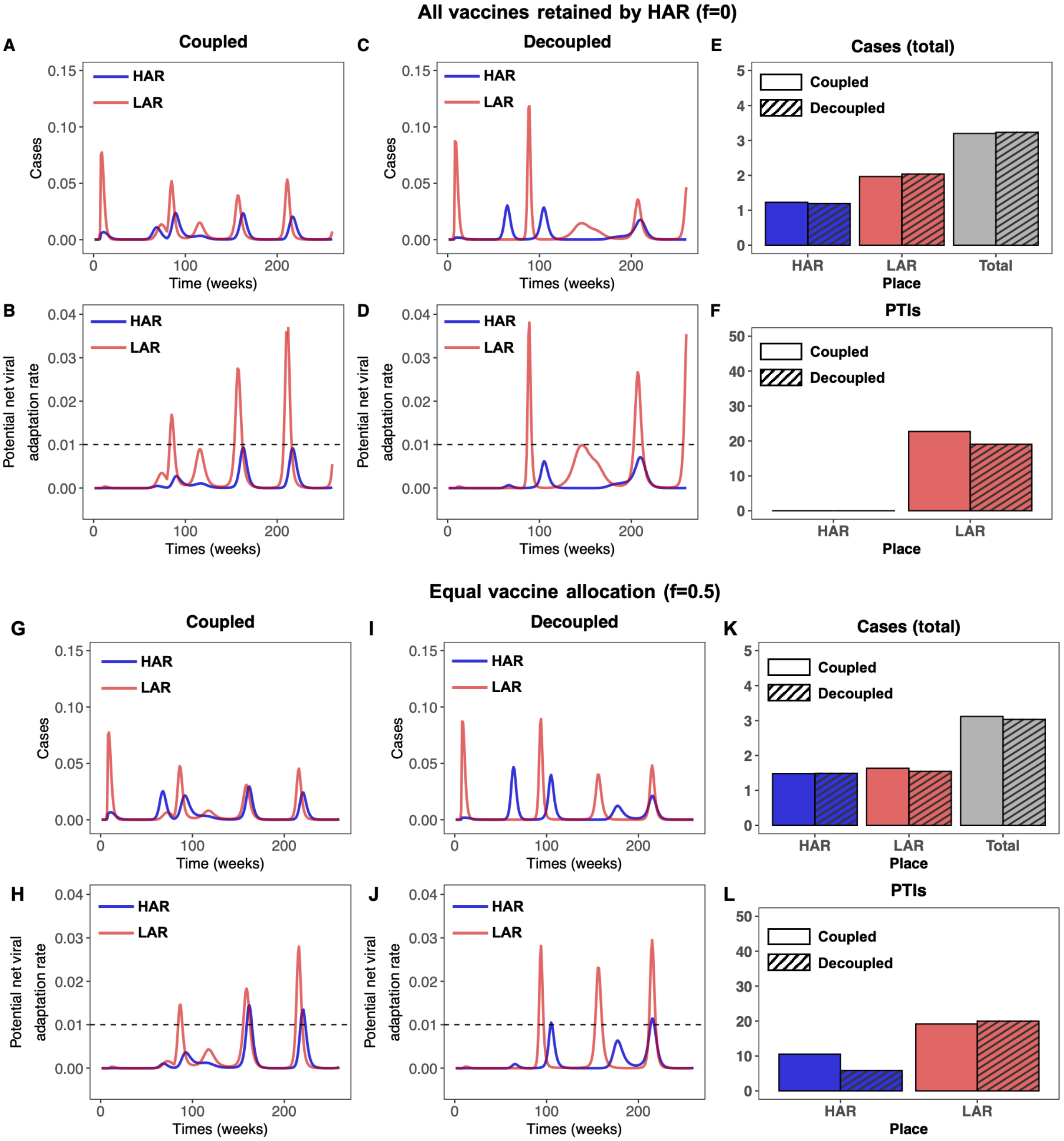

**Figure S8: Time series of cases and potential net viral adaptation rates, as well as cumulative cases and PTIs. All parameters and calculations are identical to Figure 5 of the main text, except the occurrence of a PTI is assumed to not increase the transmission rate, i.e. $\beta_i(t) = \beta_{i,0}(t)$ in both the HAR and LAR for all times (see Methods).**
